# A Novel Susceptibility Locus in *NFASC* Highlights Oligodendrocytes and Myelination in Progressive Supranuclear Palsy Pathology

**DOI:** 10.1101/2024.06.21.24309279

**Authors:** Pablo García-González, Héctor Rodrigo Lara, Yaroslau Compta, Manuel Fernandez, Sven J. van der Lee, Itziar de Rojas, Laura Saiz, Celia Painous, Ana Camara, Esteban Muñoz, Maria J. Marti, Francesc Valldeoriola, Raquel Puerta, Ignacio Illán-Gala, Javier Pagonabarraga, Oriol Dols-Icardo, Jaime Kulisevsky, Juan Fortea, Alberto Lleó, Claudia Olivé, Sterre C.M. de Boer, Marc Hulsman, Yolande A.L. Pijnenburg, Rafael Díaz Belloso, Laura Muñoz-Delgado, Dolores Buiza Rueda, Pilar Gómez-Garre, Iban Aldecoa, Gemma Aragonés, Jorge Hernandez Vara, Maite Mendioroz, Jordi Pérez-Tur, Pieter Jelle Visser, Anouk den Braber, Janne M. Papma, Ángel Martín Montes, Eloy Rodriguez-Rodriguez, Josep Blázquez-Folch, Andrea Miguel, Fernando García-Gutiérrez, Amanda Cano, Sergi Valero, Marta Marquié, María Capdevila-Bayo, Maitee Rosende-Roca, Inés Quintela, Ángel Carracedo, Lluís Tàrraga, Luis M Real, Jose Luis Royo, Maria Elena Erro, Carmen Guerrero, Daniela Corte Torres, Marta Blázquez-Estrada, Beatriz San Millán, Susana Teijeira, Dolores Vilas Rolan, Isabel Hernández, Antonio Sánchez-Soblechero, Beatriz de la Casa-Fages, Soledad Serrano López, Raquel Baviera-Muñoz, Amaya Lavín, Ricardo Taipa, Guillermo Amer, Elena Martinez-Saez, Marta Fernández-Matarrubia, Carmen Lage-Martínez, Victoria Álvarez, Laura Molina-Porcel, Henne Holstege, Pablo Mir, Olivia Belbin, Mercè Boada, Victoria Fernández, María J. Bullido, Alberto Rábano, Pascual Sánchez-Juan, Agustín Ruiz

## Abstract

We conducted the largest PSP GWAS of the Iberian population to date (522 cases from 22 Spanish and Portuguese institutions). We independently replicated seven known PSP risk variants, and unveiled a novel locus in *NFASC/CNTN2* after meta-analysing our results with a newly available Dutch cohort and publicly available summary statistics. These findings highlight the importance of neuron-oligodendrocyte interactions in PSP etiopathology.

## Main

Progressive supranuclear palsy (PSP) is a proteinopathy neuropathologically defined by intra-neuronal and intra-glial aggregations of 4-repeat microtubule associated *tau* and clinically expressed as several phenotypes with different combinations of oculomotor, parkinsonian, bulbar, cognitive, and behavioural features^1^. The common allele of an ancestral 900 kb inversion, known as the *MAPT* H1 haplotype, is the most important genetic risk factor for sporadic PSP, causing a 5.5-fold increase in PSP risk compared to the alternative *MAPT* H2 haplotype^2^. Importantly, the low prevalence of PSP conditions the scarcity of genome-wide association studies (GWAS), with only eight genetic risk loci reported to date^3–5^. Elucidating the genetic component of PSP could lead to early and improved diagnosis, identification of pathophysiological biomarkers and novel therapeutic treatments.

The PSP/DEGESCO cohort consists of clinical (N=327) and histopathological (N=195) PSP subjects from Spain and Portugal (Supplementary Table 1), genotyped using the Affymetrix 815K Spanish Biobank Array (Thermo Fisher). We leveraged our previous cohort, GR@ACE/DEGESCO^6^, to select our controls (N=1950). Since cases and controls were genotyped by the same platform and sequencing centre, we performed joint calling, quality control (QC) and TOPMed imputation^7^ of our genomic data, following standard procedures (Supplementary Information, Extended data Fig. 1). Clinical cases fulfilling the Movement Disorder Society (MDS) criteria for probable PSP^8^ were initially selected for our study, allowing the inclusion of atypical PSP phenotypes. However, we observed that atypical cases had a decreased *MAPT* H1 burden compared to those with histopathological confirmation (χ²=6.29; DF=1; *p=*0.01), and that other known PSP genetic risk factors also showed weaker associations with disease status in atypical cases compared to those with Richardson’s syndrome (PSP-RS) or histopathological confirmation (Supplementary Information), indicating the presence of phenotypic heterogeneity in our non-Richardson clinical cases (nR-PSP). As increasing the sample size in detriment of phenotypic accuracy can undermine the detection of small effect variants in GWAS studies^9^, we decided to keep only PSP-RS and histopathologically confirmed cases for further analysis.

We ran PC-adjusted case-control genome wide associations (∼9M common genetic variants) for our three subcohorts of Spanish and Portuguese ancestry (Extended Data Table 2), and meta-analyzed these results using a common effects inverse variance-weighed fixed-effect approach. We detected a strong genome-wide significant (GWS) signal (Fig. 1) for the well-known PSP-risk *MAPT* H1 haplotype (rs8070723 A: *OR[95%CI]*=4.02[3.12–5.19]; *p*=9.89·10^-27^), and replicated six additional PSP susceptibility *loci* in our study, showing consistent effect directions with previous reports (Extended Data Table 3): *MOBP* (rs1768208 G: *OR[95%CI]*=0.76[0.64– 0.89]; *p*=2.49·10^-04^)*, EIF2AK3* (rs7571971 C: *OR[95%CI]*=0.80[0.68–0.95]; *p*=9.06·10^-03^), *STX6* (rs1411478 G: *OR[95%CI]*=0.84[0.71–0.98]; *p*=0.03)*, SLCO1A2* (rs11568563 A: *OR[95%CI]*=0.54[0.42–0.69]; *p*=1.17·10^-06^), and *DUSP10 (*rs6687758 A: *OR[95%CI]*=0.80[0.66–0.97]; *p*=0.02). After adjusting by the *MAPT* H1 haplotypic background, we also reproduced an additional independent signal within the *MAPT* locus, which partially tags the PSP risk H1c subhaplotype, (rs242557 G: *OR[95%CI]*=0.72[0.61–0.86]; *p*=2.49·10^-04^). However, the effect of the GWS *RUNX2 locus,* reported by Chen *et al.*^5^, was not significant in our analysis (rs6458446 A: *OR[95%CI]*=0.90[0.75–1.09]; *p=*0.27). Additionally, a recent preprint study^10^ reported a GWS signal in the *APOE locus* where *APOE ε*4 was the protective allele and *APOE ε*2 was the risk allele. In our study, we replicated the described *APOE ε*4 protective effect (rs429358 C: *OR[95%CI]*=0.72[0.53–0.96]; *p=*0.02), but *APOE ε2* was not significant (rs7412 T: *OR[95%CI]*=1.08[0.78–1.49]; *p=*0.64). Whether a protective mechanism for *APOE ε*4, the most important genetic risk factor for sporadic AD, exists in PSP, or this effect is caused by a selection bias favouring inclusion of cases without amyloid pathology needs to be clearly stablished in future studies. Finally, the *TNXB/C4A* locus, described by another recent preprint article^11^, was not replicated (rs369580 G: *OR[95%CI]*=1.25[0.86–1.82]; *p=*0.24), although its effect direction was concordant in our cohort.

**Fig. 1.**
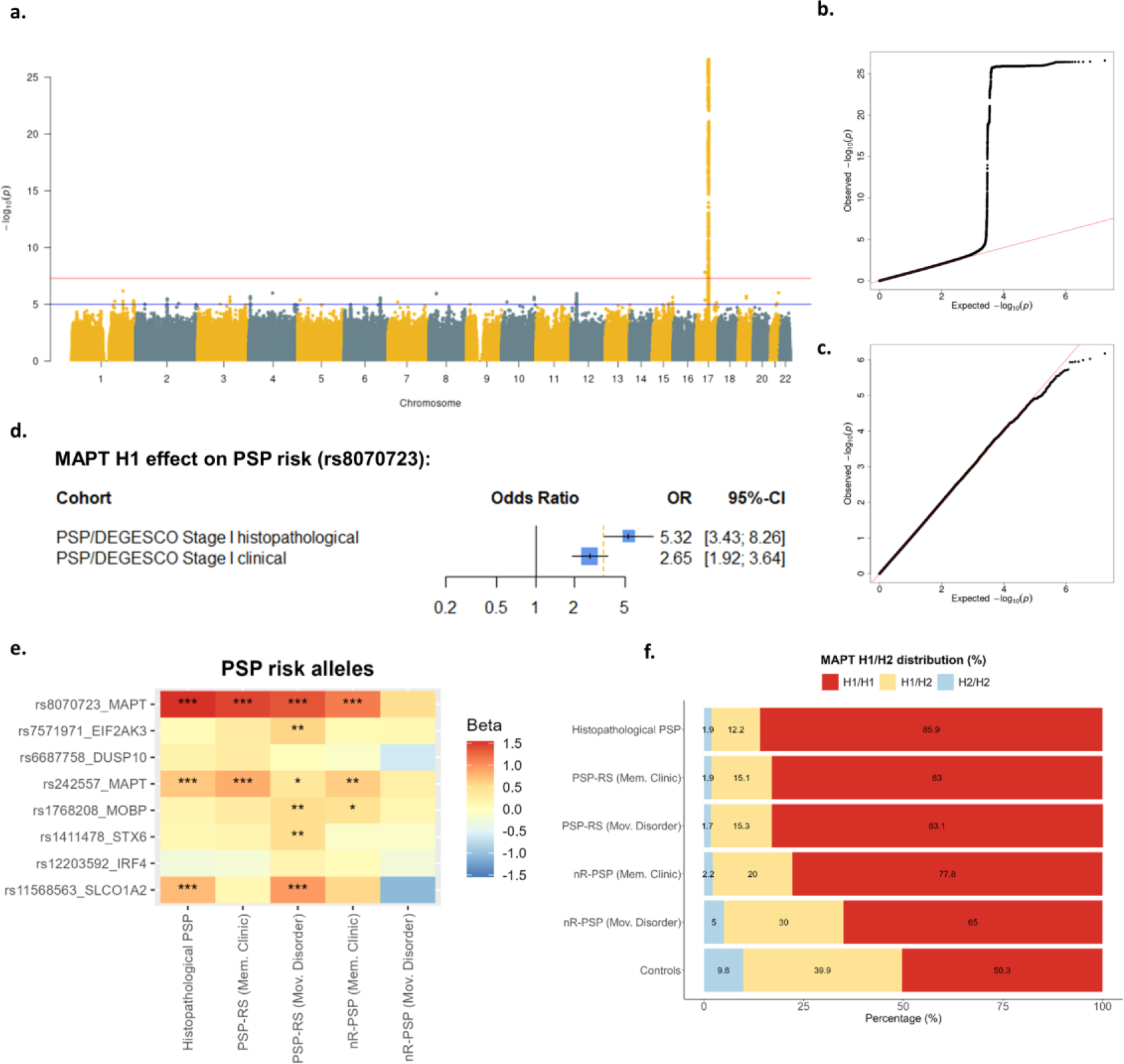
PSP GWAS in the Spanish and Portuguese population and evaluation of the clinical subgroups. **a.** Manhattan plot displaying PC-adjusted PSP genome-wide associations of common variants. **b-c.** QQ-plots displaying expected vs. observed p-values in our PSP GWAS including and excluding chr17, respectively. **d.** Forest plot displaying effects of rs8070723, a variant tagging the *MAPT* H1 haplotype in our histopathological and clinical Stage I samples. **e.** Effect sizes and significance of the risk allele of known reported PSP risk variants, as reported by Sanchez-Contreras *et al* ^4^ **f.** Distribution of *MAPT* H1/H2 haplotypes in the different groups. OR, odds ratio; PSP-RS, Richardson-Syndrome; nR-PSP, non-Richardson PSP.

Aiming to find novel PSP associations, we conducted a meta-analysis of our Spanish and Portuguese cohort with the results of the Sanchez-Contreras *et al.* study^4^, the largest publicly available summary statistics for PSP genetics, and an additional dataset including 59 clinical PSP-RS subjects and 1513 controls provided by the Amsterdam Dementia Cohort, totally comprising 3099 PSP and 11482 controls (Extended Data Table 4). An important limitation of this approach was that, due to the design of the previous studies^3,4^, only 36 SNPs were available for meta-analysis. We found a novel GWS association of the *NFASC* locus (rs12744678 C: *OR[95% CI]*=0.83[0.78–0.89]; *p*=4.15·10^-08^), which had null heterogeneity across cohorts (*I*^2^=0%), and showed concordant effect directions in all groups (Fig. 2, Extended Data Table 5). The *NFASC* gene encodes the neurofascin proteins and is highly expressed by neurons and oligodendrocytes. These proteins are involved in the development of axon initial segments and nodes of Ranvier in the central (CNS) and peripheral nervous system (PNS) by forming paranodal junctions, a very specialized cell-cell adhesion complex that is essential for correct myelination and impulse conduction^12,13^. Moreover, mutations in *NFASC* have been linked to neurodevelopmental disorders with central and peripheral motor dysfunction [61], and anti-neurofascin antibodies are found in a fraction of patients with CNS and PNS demyelinating diseases, such as multiple sclerosis^14^. Interestingly, although we did not find any significant QTLs for this variant, we found that rs4951151, the top variant on the locus (Fig. 2), is an eQTL for the neighbouring gene *CNTN2* in cerebellar tissue (on GTEx https://gtexportal.org/), and a pQTL for CNTN2 in CSF and plasma (1:205001174-T-C, on ONTIME https://ontime.wustl.edu/)15. Importantly, *CNTN2* is almost exclusively expressed by oligodendrocytes and is highly coexpressed with *MOBP* and *SLCO1A2*, two known PSP susceptibility *loci* (Extended Data Fig. 3). These three genes belong to the same expression cluster, which is implicated in myelination and enriched in this specific cell type (Extended Data Fig. 3). *CNTN2* encodes contactin-2 a transmembrane protein also implicated in the organization of nodes of Ranvier at the juxtaparanodal junctions. A study observed that the absence of this protein can impair oligodendrocyte function and axonal conduction, although it did not affect myelination under pathological conditions^16^.

**Fig. 2.**
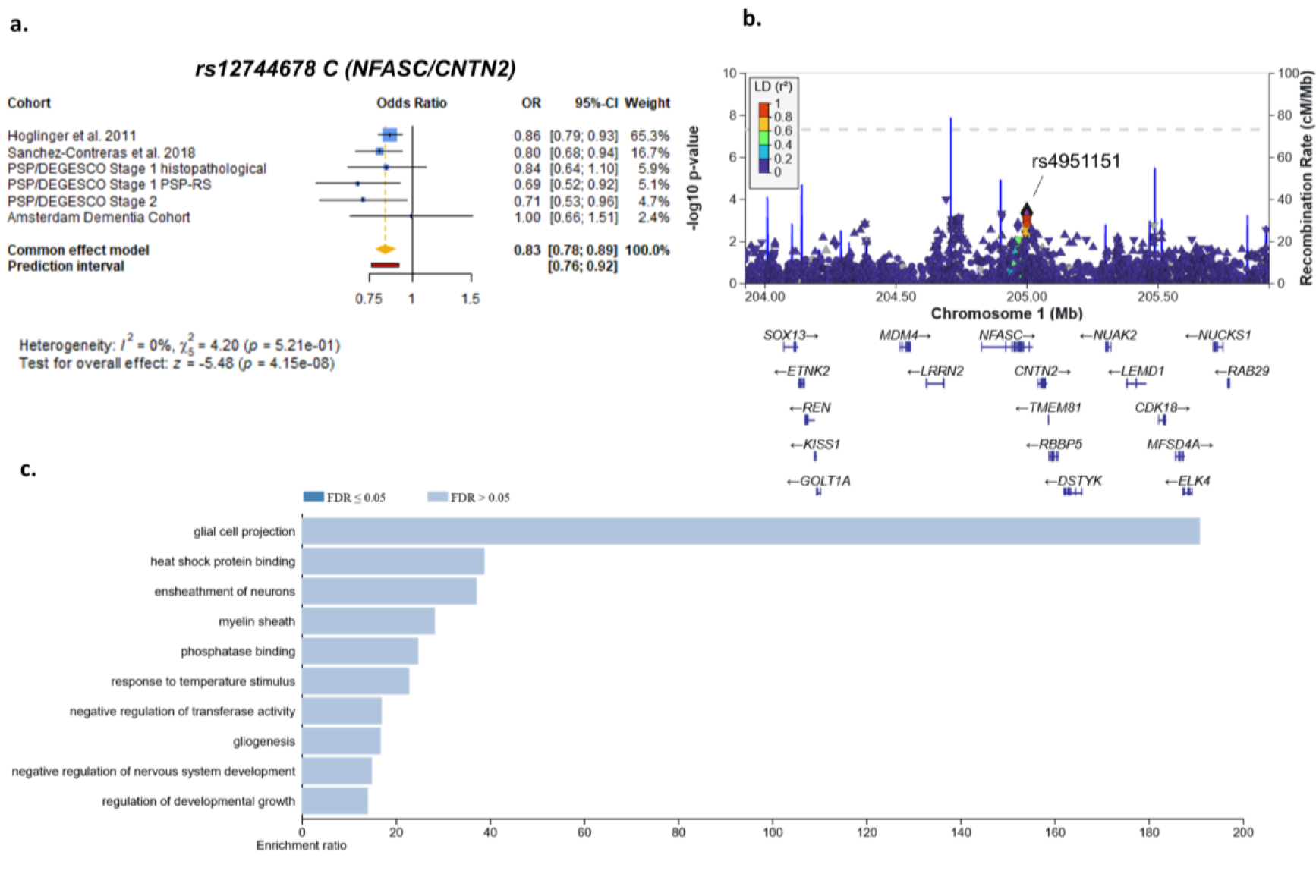
NFASC/CNTN2 is a novel locus for progressive supranuclear palsy. **a.** Forest plot displaying effect sizes and meta-analysis of the rs12744678 C allele among the different cohorts. **b.** Locus plot of the *NFASC/CNTN2* in cohorts where genome-wide association results were available (PSP/DEGESCO and ADC). **c.** Gene ontology enrichment ratio calculated using WebGestalt including PSP replicated loci (*MAPT, MOBP, EIF2AK3, STX6, DUSP10, SLCO1A2)* and the novel *NFASC* locus.

The finding of the novel *NFASC/CNTN2* locus, along with other replicated loci, such as *MOBP*, *SLCO1A2* and *DUSP10,* highlight the importance of myelination and oligodendrocyte-neuron interactions in PSP etiopathology: *MOBP* encodes the myelin-associated oligodendrocyte basic protein, a vital protein for myelin sheath structural integrity and maintenance in the central nervous system, *SLCO1A2* is highly expressed in the brain, almost exclusively by oligodendrocytes, and is highly correlated to the oligodendrocytes myelination expression cluster^17^, and *DUSP10* is highly expressed in oligodendrocytes and inhibitory neurons, encoding a phosphatase involved in MAP kinases inactivation which is able to induce oligodendrocyte differentiation (Extended Data Fig. 4)^18^. Gene ontology enrichment, including the PSP genes we replicated and the novel *NFASC* locus, reveals biological features related to myelination and axon ensheathment (Extended Data Table 6). Moreover, coexpression network analysis revealed that *MARK1*, encoding the microtubule affinity regulating kinase 1, a kinase capable of phosphorylating tau promoting microtubule destabilisation^19^, is highly coexpressed with *MOBP*, *SLCO1A2* and *NFASC* (Extended Data Fig. 5). Furthermore, a recent, preprint GWAS has also found that several PSP risk variants have oligodendrocyte-specific effects on gene expression^11^. Oligodendrocytes have a prominent microtubule structure and express *tau,* which plays a role in early contact initiation in axons, maintaining microtubule stability and myelination^20^. Moreover, tau pathology, which is predominantly neuronal in AD, is commonly found in white matter oligodendrocytes in PSP and other tauopathies^20^, and deregulation of myelination processes has been linked to neurodegenerative diseases, with a special emphasis on PSP^21^. Overall, genetic findings suggest that oligodendrocyte dysfunction and impaired myelination may play a significant role in the etiopathogenesis of PSP. Additionally, there is increasing evidence suggesting astrocytic tau pathology occurs in the earliest stages of corticobasal degeneration and PSP^22,23^. This points to a paradigm shift from neuronal to glial cell involvement in early 4-repeat tau pathology.

Of note, a small overlap may exist between our samples and some of those present in the GWAS by Höglinger *et al*^3^., as 23 cases in the former study were recruited by Spanish centres. However, this potential overlap comprises a very small fraction of the samples included in both datasets, thus we do not expect it to substantially bias our results. In fact, PSP GWASs suffer from a considerable “Russian doll” effect^9^, as new reports consist of expansions on previously available cohorts, causing a large overlap between studies. Our study used exclusively Spanish and Portuguese samples for validation of PSP susceptibility loci, and thus provides a highly independent cohort for replication of the previously reported variants (Extended Data Table 3). Importantly, there is a striking lack of diversity in PSP genetic studies to date. Expanding future studies to include non-European populations is crucial for a better understanding of the disease etiology, enhancing genetic discovery and benefiting a broader range of the human population^24^.

## Data Availability

The data that support the findings of this study are not openly available due to reasons of sensitivity and are available from the corresponding author upon reasonable request.

## Online methods

### PSP/DEGESCO cohort

Stage I samples consisted of 176 histopathologically confirmed PSP samples recruited between 2002 and 2021 from Spanish and Portuguese brain biobanks, and 312 samples fulfilling a clinical diagnosis of probable PSP provided by research groups linked to DEGESCO (Supplementary Table 1). Neuropathological diagnosis of PSP was assessed based on the distribution of neurofibrillary tangles in the brain, according to National Institute of Neurological Disorders and Stroke (NINDS) criteria^25^, while clinical diagnosis was based on the Movement Disorder Society criteria^8^. Briefly, neurologists evaluated four functional domains: ocular motor dysfunction, postural instability, akinesia and cognitive dysfunction. Considering these four domains a probable PSP diagnosis can be assigned to not only patients with PSP-RS, but also to those individuals with atypical clinical presentations, such as PSP with progressive gait freezing, with predominant parkinsonism or with predominant frontal presentation. Controls in this study were obtained from the GR@ACE/DEGESCO cohort (Supplementary Information)^6^. Finally, we recruited an additional batch of PSP samples for further validation of our results (PSP/DEGESCO Stage II), which included 39 histopathological and 154 clinical samples (Supplementary Table 1). Written informed consent was obtained from all participants in this study. This study has been approved by the competent research ethical committee (MED-FACE-2020-01, Universidad Internacional de Catalunya, Sant Cugat del Vallés, Spain).

### Amsterdam Dementia Cohort

Patient records of 397 patients diagnosed with PSP and/or corticobasal degeneration within the Amsterdam Dementia Cohort (ADC) between 2000 and 2023 were retrieved^26^. A clinician retrospectively evaluated each patient file, according the Movement Disorder Society criteria^8^, for ocular motor dysfunction, postural instability, akinesia and cognitive dysfunction. For this study, a total of 59 patients had genotyping available, were unrelated and of European ancestry, and met criteria for PSP-RS. Controls were patients without cognitive complaints from the same cohort (N=1105) and cognitively normal individuals from various other studies: one twin from each pair of twins from the EPAD study^27^ (N=97), partners of children of participants from the 100-plus Study^28^ (N=83) and controls included in the context of the Parelsnoer Institute^29^ (N=228). The local Medical Ethics Committees have approved the protocols for the ADC and the other studies and all participants of the ADC provided written informed consent.

### PSP/DEGESCO: DNA genotyping, QC & imputation

DNA from PSP cases was genotyped by the Spanish National Center for Genotyping (CEGEN) using the Axiom 815K Spanish Biobank Array (Thermo Fisher). We performed genotype calling using Affymetrix power tools (APT) software v1.15.0, and curated calls following the Axiom data analysis workflow. To improve calling accuracy we combined our cases with 5000 controls genotyped in the same center and using the same platform from the GR@ACE/DEGESCO cohort^6,30^. Called variants passing quality control (QC) criteria defined by the manufacturer (Affymetrix) were kept for downstream analysis (N=737,174). After genotype calling, we removed samples with low genotype call rates (<97%), high heterozygosity (+3SD over mean heterozygosity), discordant genetic-reported sex, and non-European ancestry, based on the 1000G European population cluster. After sample QC, we removed variants with high missingness (>5%), low frequency (MAF<0.01), differential missingness between cases and controls (p<10^-5^) or failing the Hardy Weinberg equilibrium test (p<10^-6^) in the control population. Samples with genotype call rates below 0.97 after variant QC were also removed. Additionally, we excluded duplicated and related individuals within our study by applying a GENESIS^31^ kinship filter of 0.046875. For PSP/DEGESCO Stage II samples, we merged the controls and cases passing QC from Stage I during the genotype calling stage and followed the same procedure described above. Additionally, at the IBD filtering step we removed Stage II cases overlapping or related to cases from Stage I. A complete overview of the QC process is detailed at the Supplementary Information section.

### Amsterdam Dementia Cohort: genotyping, QC and imputation

We genotyped individuals using the Illumina Global Screening Array and applied established quality control methods^32^. We used high-quality genotypes in all individuals (individual call rate >99%, variant call rate >99%), individuals with sex mismatches were excluded and departure from Hardy-Weinberg equilibrium was considered significant at p<1x10-6. Genotypes were then lifted over to GRCh38 and prepared for imputation using provided scripts (HRC-1000G-check-bim.pl) specifying TOPMed as reference panel. This script compares variant ID, strand, and allele frequencies to the TOPMed reference panel (version r2, N=194,512 haplotypes from N=97,256 individuals)^7^. Finally, all variants were submitted to the Michigan Imputation server (https://imputation.biodatacatalyst.nhlbi.nih.gov/). The server uses EAGLE (v2.4) to phase data and Minimac4 to perform genotype imputation to the reference panel. After quality control and genotype imputation of the genetic data, we kept only individuals of European ancestry (based on 1000Genomes clustering)^33^, and excluded individuals with a family relation (identity-by-descent ≥ 0.2)^34^, leaving 59 clinically diagnosed PSP-RS cases and 1513 controls for analysis.

### Statistical analysis

For GWAS analysis, we included approximately four sex-matched controls per PSP case available (Extended Data Table 2), as the increase in statistical power above this control-to-case ratio is generally negligible^35^. We removed rare SNPs (MAF<0.01) or those with low imputation quality (R2<0.3), leaving us with ∼9M variants for association testing. To test the association of SNP allele dosages with PSP risk we fitted logistic regression models using PLINK v2.00a3.7LM software. We adjusted the models by the first four PCs to account for population microstructure. As we used population-based controls in our analysis, we did not adjust the models by age to avoid jeopardizing small effects, since the large age differences between cases and controls would lead to adjustment for ‘caseness’, biasing the analysis towards the null^36^. In addition, to ensure that the association of tested variants was not age-driven, we ran PC-adjusted linear regressions for age in the full subset (N=1,450) of Stage I controls (Supplementary Table 2). Additionally, to check for H1/H2 independent associations around the *MAPT* gene, we ran models adjusted by the rs1122380 genotype, which is in full linkage disequilibrium (LD) with rs1800547 (CEU: R2=1.00; D’=1.00), the SNP previously used to report *MAPT* H1/H2 haplotype effects in our population^37^. We performed fixed effect inverse variance-weighted meta-analysis of the discovery and replication sets using the METAL software^38^. Locus association plots were made using LocusZoom (http://locuszoom.org/). All additional analyses, processing and visual representation of the data were performed using R software v4.1.1. We strand corrected and flipped effect alleles to align our results with the previous ones when needed and used the meta R package^39^ to run fixed effect inverse variance-weighed meta-analysis on specific SNPs.

### In-silico functional analysis

Enrichment analysis was performed using WebGestalt^40^ (https://www.webgestalt.org/). Expression of target genes was assessed using GTEx Portal and Human Protein Atlas for RNA and protein levels, respectively. The Online Neurodegenerative Trait Integrative Multi-Omics Explorer (ONTIME) web resource was accessed to check association of variants with quantitative endophenotypes^15^. Finally, gene co-expression networks were generated using GeneFriends (https://www.genefriends.org/)41. Details on these methodologies are provided in the Supplementary Information.

## Data availability

The data that support the findings of this study are not openly available due to reasons of sensitivity and are available from the corresponding author upon reasonable request. Summary statistics detailing association results will be made publicly available upon acceptance of the manuscript.

## Code availability

Custom code was used to calculate polygenic risk scores and is publicly available (https://github.com/Pablo-GarGon/PRS_Generator). Other analyses were performed using publicly available software, tools and algorithms, as detailed in the Methods and Supplementary Information sections.

## Acknowledgements

The present work has been performed as part of the Doctoral thesis of Pablo García-González at the University of Barcelona (Barcelona, Spain). We would like to thank patients and controls who participated in this project. Some control samples and data from patients included in this study were provided in part by the National DNA Bank Carlos III (www.bancoadn.org, University of Salamanca, Spain) and Hospital Universitario Virgen de Valme (Sevilla, Spain); they were processed following standard operating procedures with the appropriate approval of the Ethical and Scientific Committee. We are grateful to the Biomedic Diagnostic Center (CBD) of the Hospital Clinic de Barcelona for access to, and assistance with, FLUOSTAR Omega equipment and RT-QuIC experiments (Drs. Raquel Ruiz and Laura Naranco). Hospital Clinic de Barcelona receives support from the CERCA programme from Generalitat de Catalunya (Catalan government). Several authors of this manuscript are members of the European Reference Network for Rare Neurological Diseases - Project ID No 739510, as well as CIBERNED (CB06/05/0018-ISCIII). We acknowledge the Biobank of the University of Navarra for its collaboration. We thank all study participants and all personnel involved in data collection for the contributing studies. Research of Alzheimer Center Amsterdam is part of the neurodegeneration research program of Amsterdam Neuroscience. Alzheimer Center Amsterdam is supported by Stichting Alzheimer Nederland and Stichting Steun Alzheimercentrum Amsterdam. The chair of Wiesje van der Flier is supported by the Pasman stichting. The clinical database structure was developed with funding from Stichting Dioraphte. The work in this manuscript was carried out on the Snellius supercomputer, which is embedded in the Dutch national e-infrastructure with the support of SURF Cooperative. Computing hours were granted in 2016, 2017, 2018 and 2019 to H. Holstege by the Dutch Research Council (project name: ‘100plus’; project numbers 15318 and 17232). Samples and data from patients included in this study were provided by the Vall d’Hebron University Hospital Biobank (PT20/00107), integrated in the Spanish National Biobanks Network, and they were processed following standard operating procedures with the appropriate approval of the Ethical and Scientific Committees. We are indebted to the Biobank-Hospital Clinic-FRCB-IDIBAPS for samples and data procurement.This work was funded by CIBERNED (Centro Investigación Biomédica en Red Enfermedades Neurodegenerativas) 10th internal call for cooperative projects (Register number: 2019/09). Agustín Ruiz, Pascual Sanchez-Juan, Alberto Rábano, and Maria J. Bullido received funding from this convocatory. Pablo García-González received support by CIBERNED employment plan CNV-304-PRF-866. CIBERNED is integrated into ISCIII (Instituto de Salud Carlos III). Mercé Boada, Marta Marquié and Agustín Ruiz are also supported by national grants PI13/02434, PI16/01861, PI17/01474, PI19/01240. PI19/01301, PI19/00335 and PI22/011403, and CIBERNED grant 2019/08. The Genome Research @ Fundació ACE project (GR@ACE) is supported by Grifols SA, Fundación bancaria “La Caixa”, Fundació ACE, and CIBERNED. Acción Estratégica en Salud is integrated into the Spanish National R + D + I Plan and funded by ISCIII—Subdirección General de Evaluación and the Fondo Europeo de Desarrollo Regional (FEDER—“Una manera de hacer Europa”). Agustín Ruiz is supported by ISCIII national grant PMP22/00022, funded by the European Union (NextGenerationEU). The genotyping service to generate GR@ACE and PSP/DEGESCO GWAS data was carried out at CEGEN-PRB3-ISCIII; it is supported by grant PT17/0019, of the PE I+D+i 2013-2016, funded by ISCIII and ERDF”. Itziar de Rojas received support by a national grant from ISCIII FI20/00215. Amanda Cano acknowledges the support of the Spanish Ministry of Science, Innovation and Universities under the grant Juan de la Cierva (FJC2018-036012-I), the support of ISCIII under the grant Sara Borrell (CD22/00125), Spanish Ministry of Science and Innovation, Proyectos de Generación de Conocimiento grant PID2021-122473OA-I00, and Fundación ADEY under the program “Proyectos de Investigación en Salud 2023”. Fondo de Investigacion en Salud (Instituto Carlos III) (PI17/00096) in the frame of the European Regional Development Fund (ERDF) and to Fundacio la Marato de TV3 (PI043296 and 202009-10). This work was supported by the Spanish Ministry of Science and Innovation (RTC2019-007150-1), the Instituto de Salud Carlos III (ISCIII) and co-funded by the European Union (PI14/01823, PI16/01575, PI18/01898, PI19/01576, PI21/01875), the Consejería de Economía, Innovación, Ciencia y Empleo de la Junta de Andalucía (CVI-02526, CTS-7685, PY20_00896), and the Consejería de Salud y Bienestar Social de la Junta de Andalucía (PI-0471-2013, PE-0210-2018, PI-0459-2018, PE-0186-2019). Pilar Gómez-Garre was supported by the “Nicolás Monardes” program (C-0048-2017) from Andalusian Regional Ministry of Health. Laura Muñoz-Delgado was supported by the “Río Hortega” program [CM21/00051] from the Instituto de Salud Carlos III (ISCIII-FEDER). The funders had no role in study design, data collection and analysis, decision to publish, or preparation of the manuscript. Sven J. van der Lee was funded for this study by NWO (#733050512, PROMO-GENODE: a PROspective study of MOnoGEnic causes Of Dementia) a substantial donation by Edwin Bouw Fonds and Dioraphte. SvdL further received funding for the GeneMINDS consortium, which is powered by Health∼Holland, Top Sector Life Sciences & Health. Sven J. van der Lee is a recipient of ABOARD, which is a public-private partnership receiving funding from ZonMW (#73305095007) and Health∼Holland, Topsector Life Sciences & Health (PPP-allowance; #LSHM20106). More than 30 partners participate in ABOARD. ABOARD also receives funding from Edwin Bouw Fonds and Gieskes-Strijbisfonds. ABOARD also receives funding from de Hersenstichting, Edwin Bouw Fonds and Gieskes-Strijbisfonds. Array genotyping was performed in the context of EADB (European Alzheimer DNA biobank) funded by the JPco-fuND FP-829-029 (ZonMW projectnumber 733051061). This work has been supported by the Queen Sofia Foundation. Pascual Sánchez-Juan is supported by grants from ISCIII (PMP22/00022 and PI20/01011) and TED2021-131676B-100. Luis Miguel Real received support from “The absence of seroconversion after exposition to hepatitis C virus is not related to KIR-HLA genotype combinations” (GEHEP-012 study). Consejería de Salud de la Junta de Andalucía (grant number PI-0001/2017) and CIBERINFEC -Consorcio Centro de Investigación Biomédica en Red de Enfermedades Infecciosas-Instituto de Salud Carlos III, Ministerio de Ciencia e Innovación y Unión Europea (Spain) (NextGeneration EU) (grant number CB21/13/00118). Jose Luis Royo received support from Project 23.04.2021 from Santángela Foundation (Sevilla, Spain).

## Contributions

P.Gar-Gon. wrote the manuscript. Y.C., V.F., M.Bu., A.Ra., P.S-J., and A.Ru. jointly supervised the work. H.R.L., M.Bu., A.Ra., P.S-J., and A.Ru. designed and conceptualized the study. P.Gar-Gon., S.vdL., and M.H performed bioinformatics and statistical analyses. P.Gar-Gon., M.F., S.vdL., I.dR., L.S., R.P., C.O., Y.P., I.Q., A.C., and H.H. were involved in data generation. H.R.L., Y.C., M.F., S.vdL., L.S., C.P., A.Cam., E.M., M.J.M., F.V., I. I-G., J.P., I. D-I., J, K, J.F., A.Ll., S.CM.dB., Y.P., R.D-B., L.M-D., D.B-R., P.Gom-Gar., I.A., G.Ar., J.H.V., M.Me., J.P-T-, P.J.V., A.d.B., J.M.P., A.M.M., L.T., L.M.R., JL. R., M.E.E., C.G., D.C.T., M.B.E., B.S.M., S.T.B., D.V.R., I.H., A.S-S., B.C-F., S.R.L., R.B.M., A.La., R.T., G.Am., E.M.S., M.F-M., C.L-M., V.A., L.M.P., H.H., P.M., O.B., and M.Bo. were involved in sample contribution. All authors critically revised the manuscript for important intellectual content and approved this final version.

## Extended Data

**Extended Data Table 1.** Summary of samples remaining after QC steps in the PSP/DEGESCO cohort.

| *PSP/DEGESCO Stage I* |  | **PSP** |  | **Controls** |  |  |
| --- | --- | --- | --- | --- | --- | --- |
| **QC step:** | **N** | **Fail** | **Fail (%)** | **N** | **Fail** | **Fail (%)** |
| **DNA QC** | 488 | 88 | 18.03% | NA | NA | NA |
| **Calling** | 400 | 8 | 2.00% | 5000 | 26 | 0.52% |
| **Heterozygosity** | 392 | 3 | 0.77% | 4974 | 4 | 0.08% |
| **Sexcheck** | 389 | 10 | 2.57% | 4970 | 0 | 0.00% |
| **PCA** | 379 | 6 | 1.58% | 4970 | 15 | 0.30% |
| **IBD** | 373 | 20 | 5.36% | 4955 | 300 | 6.05% |
| **FINAL** | **353** |  |  | **4655** |  |  |
| *PSP/DEGESCO Stage II* |  | **PSP** |  | **Controls** |  |  |
| **QC step:** | **N** | **Fail** | **Fail (%)** | **N** | **Fail** | **Fail (%)** |
| **DNA QC** | 193 | 0 | 0.00% | NA | NA | NA |
| **Calling** | 193 | 12 | 6.22% | 5000 | 12 | 0.24% |
| **Heterozygosity** | 181 | 0 | 0.00% | 4988 | 0 | 0.00% |
| **Sexcheck** | 181 | 1 | 0.55% | 4988 | 0 | 0.00% |
| **PCA** | 180 | 0 | 0.00% | 4988 | 10 | 0.20% |
| **IBD** | 180 | 11 | 6.11% | 4978 | 302 | 6.07% |
| **FINAL** | **169** |  |  | **4676** |  |  |
| N, sample size |  |  |  |  |  |  |

**Extended Data Table 2.** Demographic features of samples included in the Spanish and Portuguese PSP GWAS used for replication of previous known PSP *loci*.

|  | CA |  |  | CO |  |  |
| --- | --- | --- | --- | --- | --- | --- |
|  | N | %Males | Age* (SD) | N | %Males | Age* (SD) |
| **PSP/DEGESCO Stage I (histopathological)** | 156 | 56.4 | 76.5 (9.5) | 650 | 56.5 | 67.8 (10.1) |
| **PSP/DEGESCO Stage I (PSP-RS)** | 112 | 48.2 | 67.2 (7.8) | 800 | 46.8 | 68.6 (10.3) |
| **PSP/DEGESCO Stage II (hist. + PSP-RS)** | 119 | 54.6 | 69.2 (8.2) | 500 | 54.6 | 68.1 (10.5) |
| **TOTAL** | 465 |  |  | 1950 |  |  |

N, sample size; SD, standard deviation. \*Controls: Mean age at last visit; histopathological PSP: Mean age at death; clinical PSP: Mean age at motor symptoms onset.

**Extended Data Table 3.**
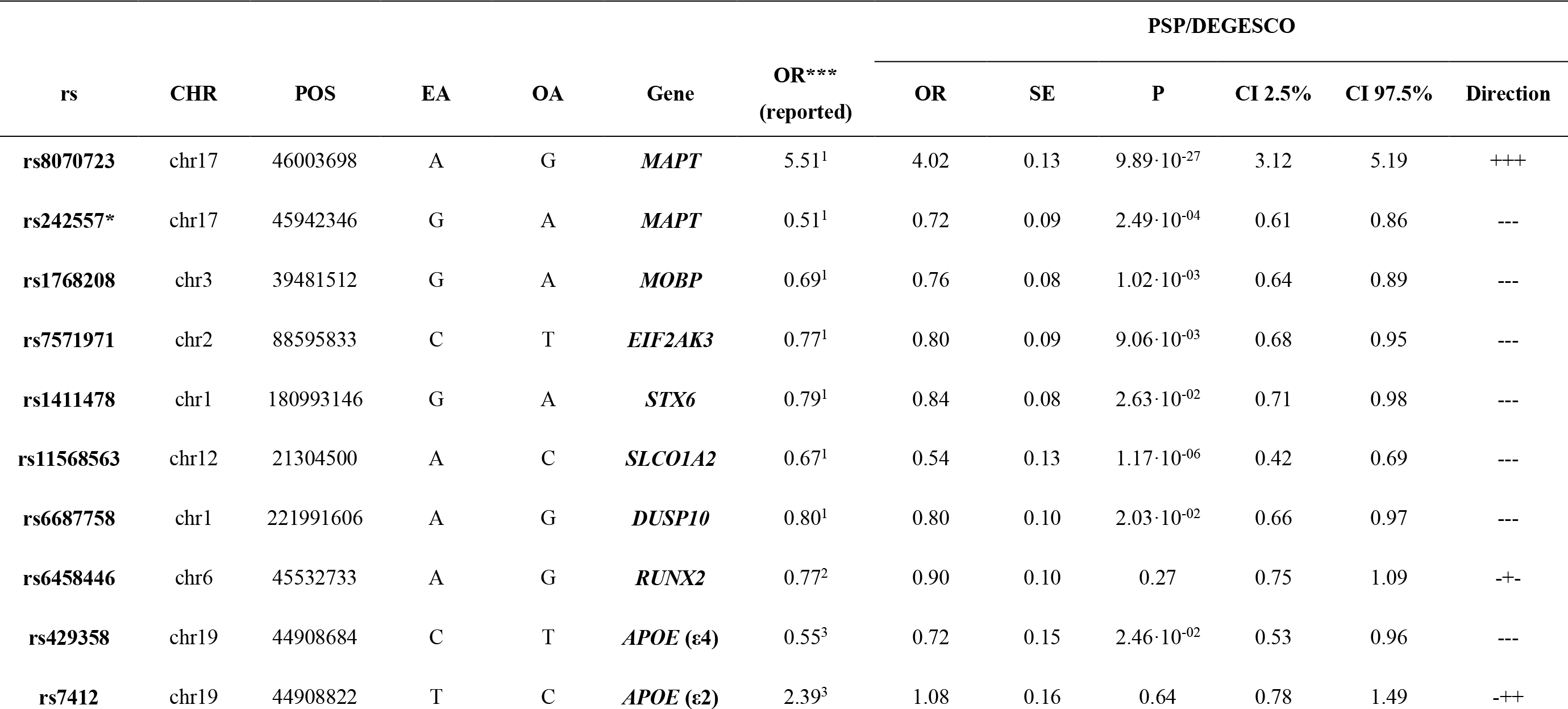
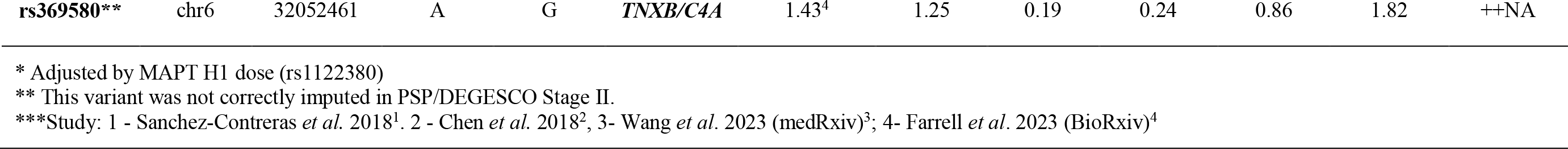
Summary statistics of the replication in Spanish and Portuguese population of PSP GWS susceptibility *loci* reported to date. The direction column shows the effect directions in the PSP/DEGESCO Stage I histopathological, PSP/DEGESCO Stage I PSP-RS, and PSP/DEGESCO Stage II, respectively. Genomic coordinates correspond to the GRCh38 assembly. EA, effect allele; OA, other allele; OR, odds ratio; SE, standard error; P, p-value; CI, confidence interval.

**Extended Data Table 4.** Main features of the cohorts available for meta-analysis.

| Study | Variants<br>available | PSP |  |  | Control |  |  |
| --- | --- | --- | --- | --- | --- | --- | --- |
|  |  | N | Males (%) | Age (SD) | N | Males (%) | Age (SD) |
| Höglinger *et al.* 2011<sup>5</sup> | 4,099 | 2120 | 54.0 | ~66.5 (NA) | 6847 | NA | NA |
| Sanchez-Contreras *et al.* 2018<sup>1</sup> | 37 | 533 | 55.0 | 75.0 (7.6) | 1172 | 52.5 | 77.0 (8.9) |
| PSP/DEGESCO Stage I (histopathological) | 8,990,045 | 156 | 56.4 | 76.5 (9.5) | 650 | 56.5 | 67.8 (10.1) |
| PSP/DEGESCO Stage I (PSP-RS) | 9,008,824 | 112 | 48.2 | 67.2 (7.8) | 800 | 46.8 | 68.6 (10.3) |
| PSP/DEGESCO Stage II (hist. + PSP-RS) | 8,433,148 | 119 | 54.6 | 69.2 (8.2) | 500 | 54.6 | 68.1 (10.5) |
| Amsterdam Dementia Cohort (PSP-RS) | 8,794,028 | 59 | 54.2 | 67.7 (5.5) | 1513 | 58.5 | 62.3 (10.5) |
| Meta-analysis | 36 | 3099 |  |  | 11482 |  |  |

N, sample size; SD, standard deviation.
\*Controls: Mean age at last visit; histopathological PSP: Mean age at death; clinical PSP: Mean age at motor symptoms onset.

**Extended Data Table 5.** Summary statistics of the meta-analysis of the 36 SNPs available SNPs including data from the Höglinger *et al*. _5_ **and Sanchez-Contreras et al.**_1_ **studies, PSP/DEGESCO and ADC.** Genomic coordinates correspond to the GRCh38 assembly. EA, effect allele; OA, other allele; OR, odds ratio; P, p-value; EAF, effect allele frequency; SE, standard error; CI, confidence interval; I2, heterogeneity estimate. ***UPPLIED SEPARATELY IN EXCEL FORMAT\**** ***\*SUPPLIED SEPARATELY IN EXCEL FORMAT\****

**Extended Data Table 6.** Top enriched genesets using the WebGestalt Over Representation Analysis including the 6 PSP loci replicated in the PSP/DEGESCO cohort (*MAPT, MOBP, EIF2AK3, STX6, DUSP10* and *SLCO1A2*) and the novel *NFASC* locus. Size depicts the number of genes in the geneset. Overlap depicts the number of genes in our input list present in the geneset. Expect depicts the number of genes expected to overlap with each geneset at random given the number of imputed genes.

| geneSet | Description | Size | Overlap | Expect | Enrichment<br>Ratio | P | FDR | database |
| --- | --- | --- | --- | --- | --- | --- | --- | --- |
| GO:0097386 | glial cell projection | 23 | 2 | 0.01047563 | 190.9 | 4.48E-05 | 0.06 | Cellular Component |
| GO:0031072 | heat shock protein binding | 113 | 2 | 0.05146724 | 38.9 | 1.10E-03 | 0.52 | Molecular Function |
| GO:0007272 | ensheathment of neurons | 118 | 2 | 0.05374455 | 37.2 | 1.20E-03 | 0.52 | Biological Process |
| GO:0043209 | myelin sheath | 155 | 2 | 0.07059666 | 28.3 | 2.05E-03 | 0.67 | Cellular Component |
| GO:0019902 | phosphatase binding | 177 | 2 | 0.08061683 | 24.8 | 2.67E-03 | 0.68 | Molecular Function |
| GO:0009266 | response to temperature stimulus | 192 | 2 | 0.08744876 | 22.9 | 3.13E-03 | 0.68 | Biological Process |
| GO:0051348 | negative regulation of transferase activity | 258 | 2 | 0.11750927 | 17.0 | 5.58E-03 | 0.69 | Biological Process |
| GO:0042063 | gliogenesis | 261 | 2 | 0.11887566 | 16.8 | 5.70E-03 | 0.69 | Biological Process |
| GO:0051961 | negative regulation of nervous system development | 294 | 2 | 0.13390591 | 14.9 | 7.19E-03 | 0.69 | Biological Process |
| GO:0048638 | regulation of developmental growth | 312 | 2 | 0.14210424 | 14.1 | 8.06E-03 | 0.69 | Biological Process |
| P, p-value; FDR, false discovery rate adjusted p-value. |  |  |  |  |  |  |  |  |

**Extended Data Fig. 1.**
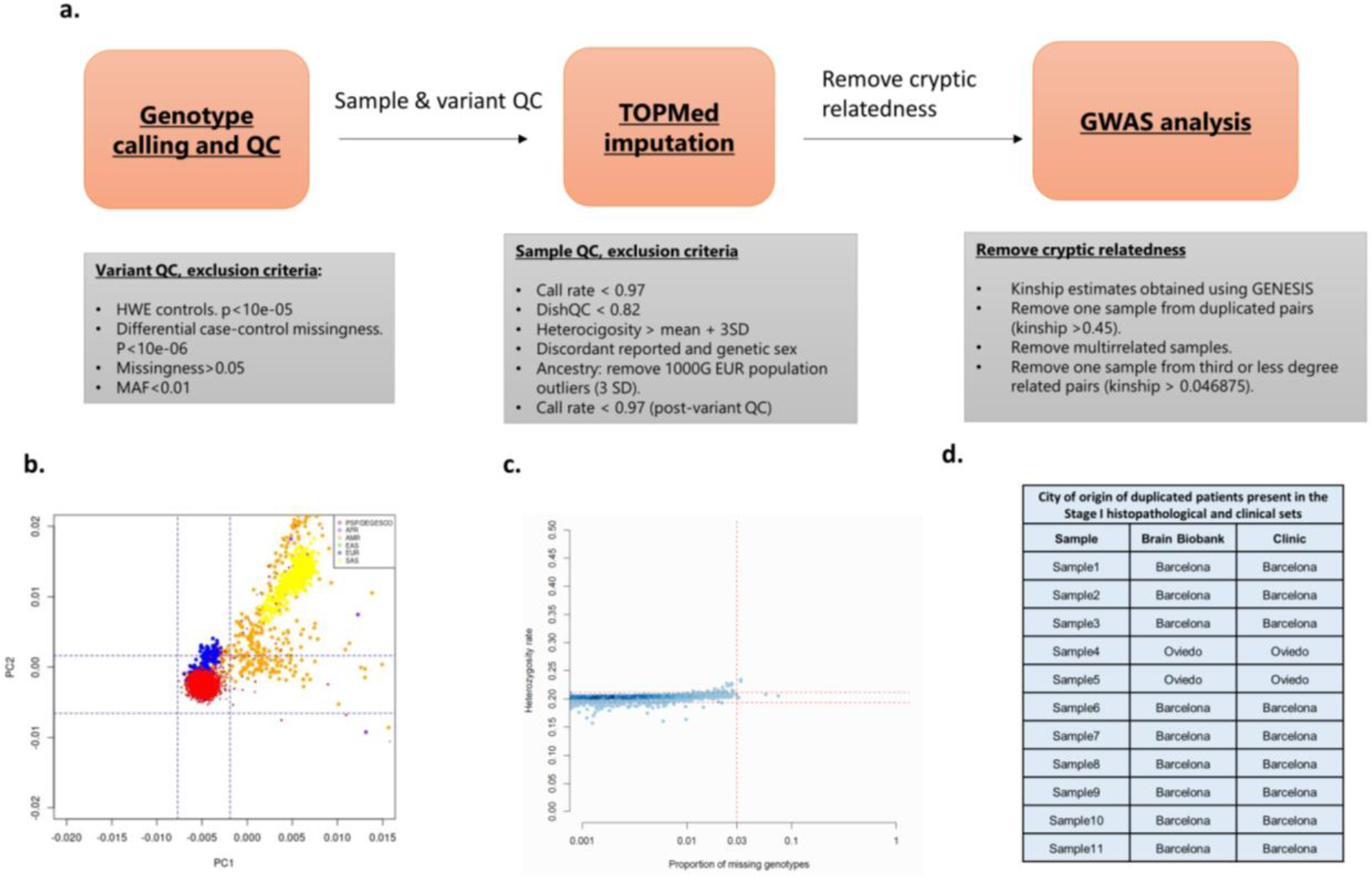
**a.** Overview of the quality control process for PSP cases and controls. **b.** Scatterplot of the first two principal components calculated using the 1000 Genomes dataset^6^, showing the different 1000G populations and the samples in PSP/DEGESCO Stage I. We removed samples in PSP/DEGESCO that were outliers (3 standard deviations away from the mean) with respect to the 1000G European cluster. **c.** Scatterplot representing sample call rates and heterozygosity rates in PSP/DEGESCO Stage I. Contaminated samples are expected to display an increase in both the proportion of uncalled genotypes and heterozygosity rate. **d**. City of origin of duplicated individuals found across PSP/DEGESCO Stage I clinics and brain biobanks.

**Extended Data Fig. 2.**
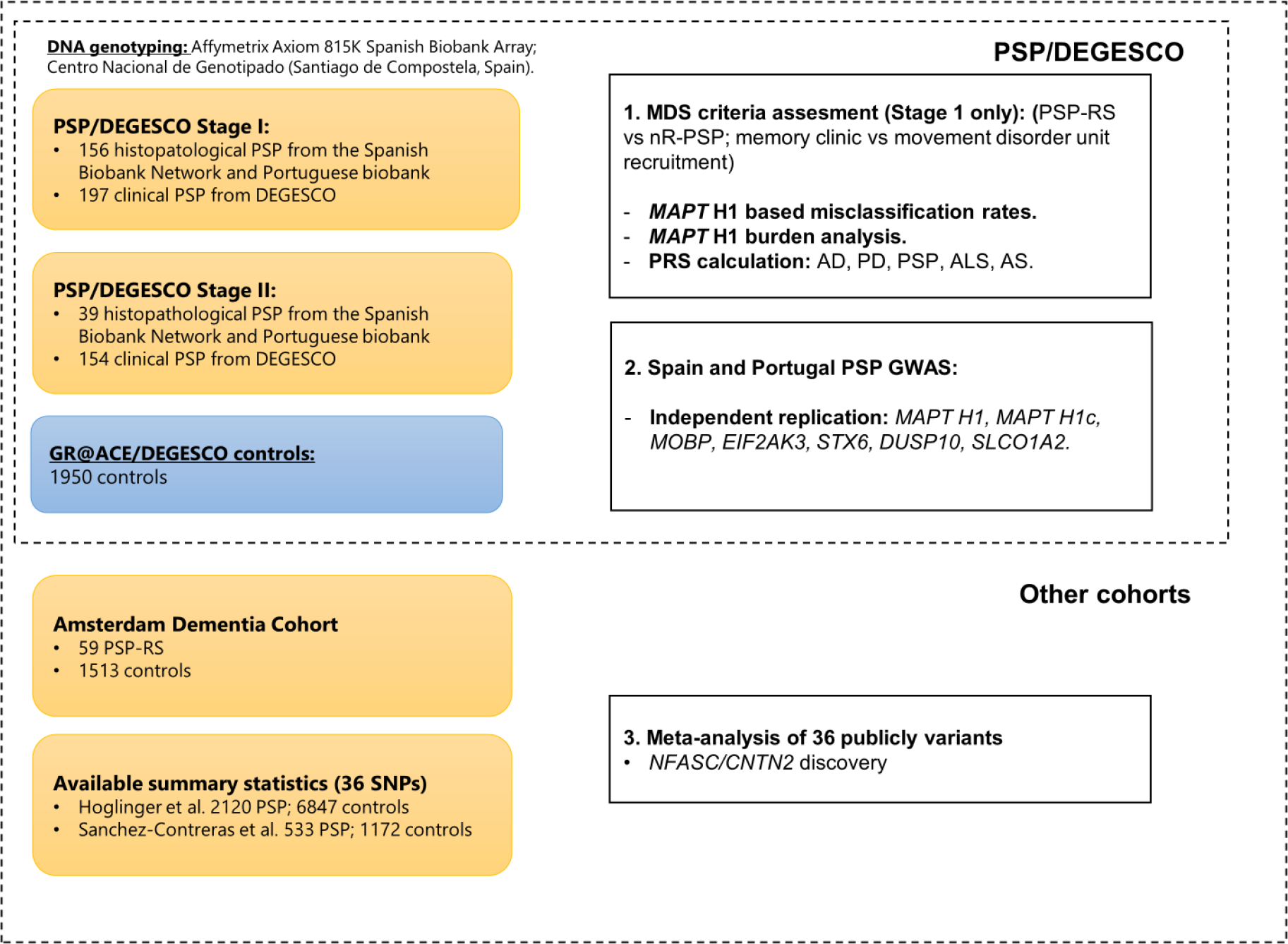
Schematic depiction of cohorts used and steps taken in our study.

**Extended Data Fig. 3.**
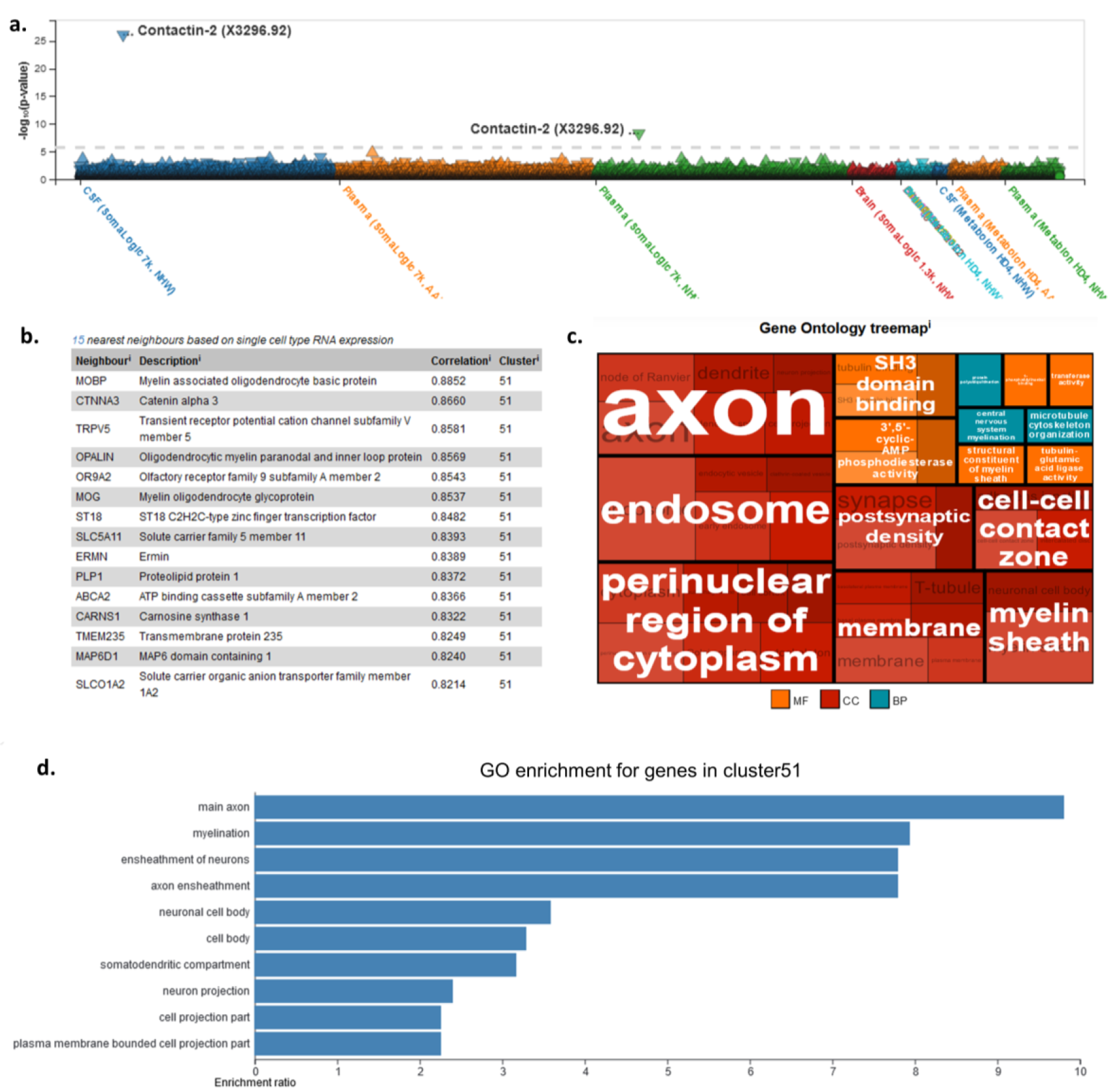
**a.** Association of rs4951151, the top variant detected in the *NFASC/CNTN2* locus with quantitative endophenotypes available on ONTIME https://ontime.wustl.edu/)7. Significant signals were observed for contactin-2 protein levels in CSF and plasma. **b.** Top correlated genes in the Human Protein Atlas single cell expression cluster^8^ for CNTN2, referred to as ‘cluster 51’. **c.** Gene ontology treemap of the expression cluster 51, showing the main molecular functions, cellular components and biological processes enriched among these genes. **d.** Gene ontology enrichment for genes in cluster 51 performed using WebGestalt^9^.

**Extended data Fig. 4.**
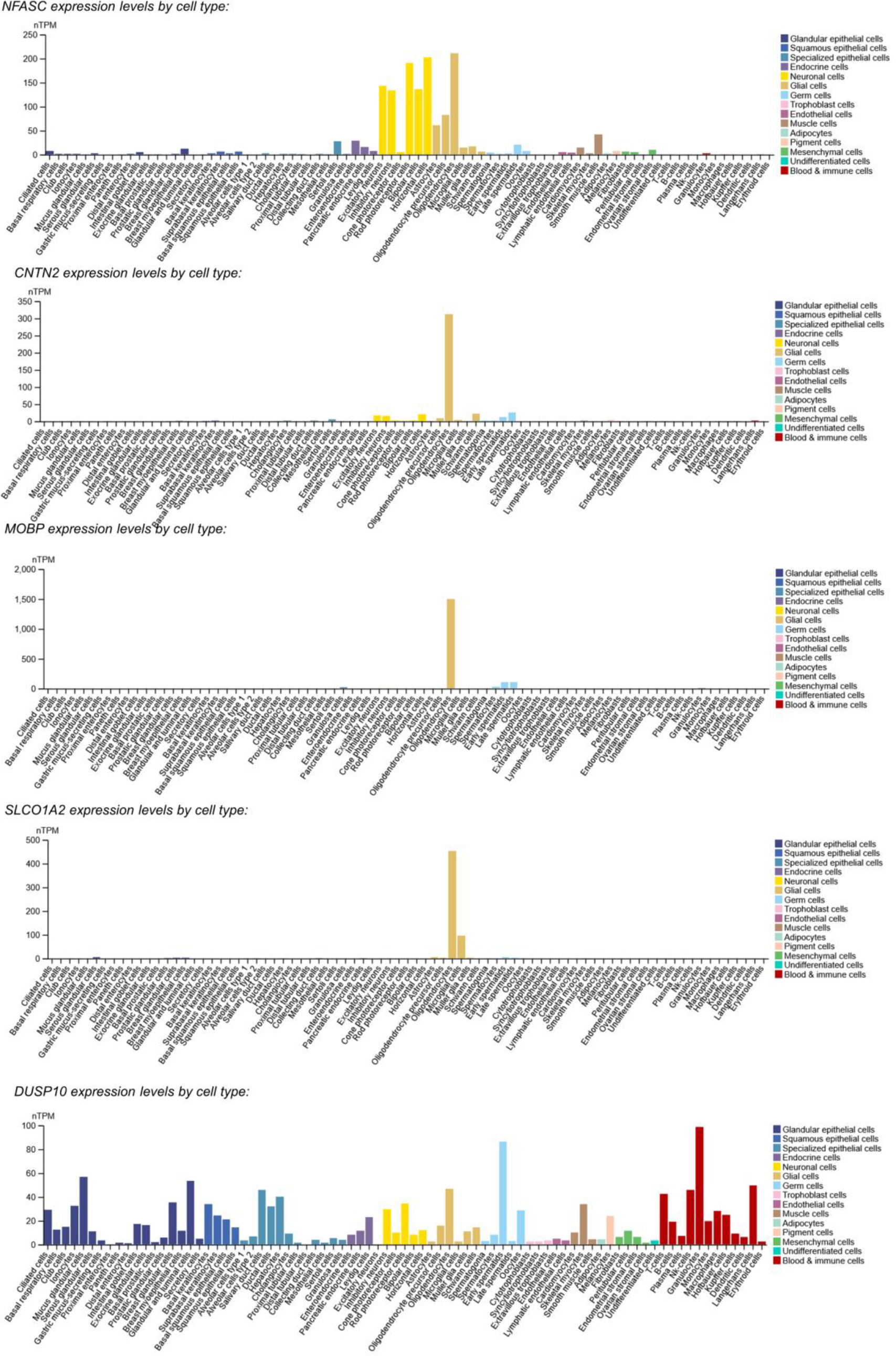
RNA expression levels by cell type for *MOBP*, *SLCO1A2*, *DUSP10*, *NFASC* and *CNTN2,* respectively, based on data from the Human Protein Atlas^8^.

**Extended Data Fig. 5.**
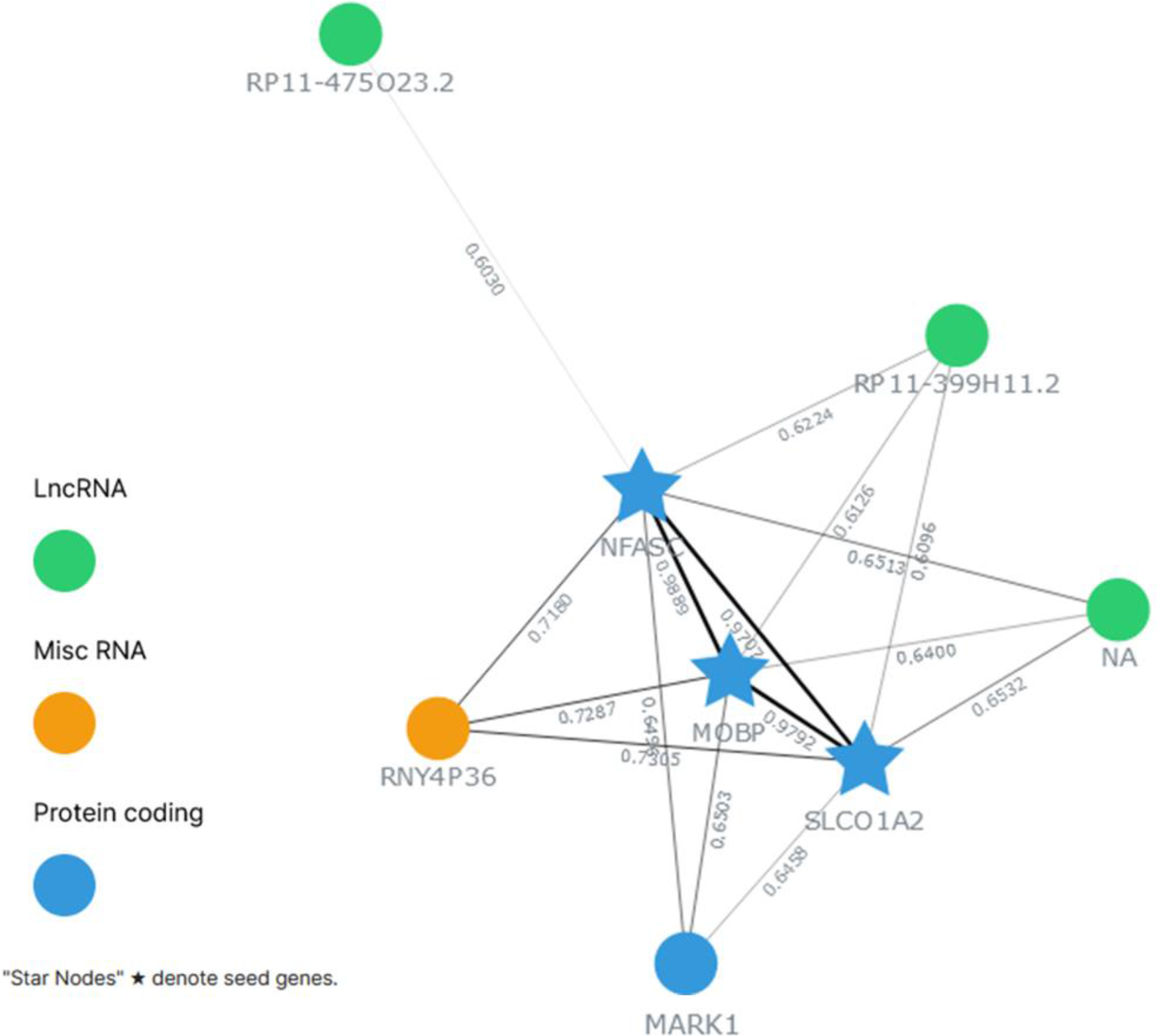
Gene coexpression network generated using GeneFriends^10^ from our input gene list: *MAPT, MOBP, EIF2AK3, STX6, DUSP10, SLCO1A2, NFASC* and *CNTN2.* Edges were labelled with the Pearson correlation coefficient existent between transcripts belonging to the network.

## Supplementary Information: Supplementary Information: A Novel Susceptibility Locus in NFASC Highlights Oligodendrocytes and Myelination in Progressive Supranuclear Palsy Pathology

### 1. Supplementary notes: DNA genotyping, QC & imputation in the PSP/DEGESCO cohort

DNA from PSP cases was extracted from 25 mg of cerebellar cortex (histopathological PSP) or peripheral blood (clinical PSP). After excluding samples with insufficient concentration (<5 ng/µL) or degraded DNA, samples were genotyped by the Spanish National Center for Genotyping (CEGEN) using the Axiom 815K Spanish Biobank Array (Thermo Fisher). We performed genotype calling and QC using Affymetrix power tools (APT) software v1.15.0 following the Axiom data analysis workflow. We kept samples with dishQC values >0.82, a parameter determined by measuring the resolution of AT and GC channels within monomorphic SNPs, and genotype call rates >0.97. We also checked that the average call rate within plates was >0.985 to detect potentially problematic batches. We jointly performed genotype calling for the remaining PSP samples and 5,000 Spanish controls obtained from the GR@ACE/DEGESCO cohort, which had been previously genotyped with the same platform, using the same array and in the same centre, thus minimizing potential batch effects^1,2^. Called variants passing quality control (QC) criteria defined by the manufacturer (Affymetrix) were kept for downstream analysis (N=737,174).

After genotype calling, we removed samples with low genotype call rates (<97%) and high heterozygosity (+3SD over mean heterozygosity). To identify mislabelled samples, we performed a sex-check and removed those with discordant reported and genetic sex. We ran a principal component (PC) analysis to identify and remove samples from non-European ancestries based on the 1000 Genomes European population cluster (Extended Data Fig. 1). After sample QC, we removed variants with high missingness (>5%), low frequency (MAF<0.01), differential missingness between cases and controls (p<10^-5^) or failing the Hardy Weinberg equilibrium test (p<10^-6^) in the control population. Samples with genotype call rates below 0.97 after variant QC were also removed from the dataset. Genotype imputation was performed for 379 PSP samples and 4,955 controls in the TOPMed Imputation Server (Michigan, USA) using the TOPMed reference panel based on 587,195 genotyped variants passing QC. Prior to GWAS analysis, we used the GENESIS R package^3^ to detect and remove potential cryptic relatedness within our population. We first used a kinship filter of 0.45 to identify and remove sample pairs corresponding to the same individual (N=11). After removing duplicates, we detected related pairs by applying a kinship filter of 0.046875, which represents the theoretical kinship value between third-and fourth-degree relatedness. We first removed the samples present in multiple related pairs, and for the remaining unique relative pairs we applied the following criteria: 1) if one sample was a PSP and the other one was a control we removed the control; 2) if one sample was a histopatologically confirmed PSP the other sample was a clinical PSP, we kept the histopathological PSP; 3) if both samples belonged to the same clinical group (control, clinical or histopathological PSP), we kept the sample with the highest genotype call rate. For PSP/DEGESCO Stage II samples, we performed the genotype calling including the controls and cases passing QC from Stage I, and followed the same procedure described above. Additionally, at the IBD filtering step we removed Stage II cases overlapping or related to cases from Stage I. An overview of the complete QC process is provided (Extended Data Fig. 1, Extended Data Table 1, Supplementary Table 1).

### 2. Supplementary notes: DNA genotyping, QC & imputation in the Amsterdam Dementia Cohort

We genotyped all individuals using the Illumina Global Screening Array and applied established quality control methods^4^. We used high-quality genotypes in all individuals (individual call rate >99%, variant call rate >99%), individuals with sex mismatches were excluded and departure from Hardy-Weinberg equilibrium was considered significant at p<1x10-6. Genotypes were then lifted over to GRCh38 and prepared for imputation using provided scripts (HRC-1000G-check-bim.pl) specifying TOPMED as reference panel. This script compares variant ID, strand, and allele frequencies to the TOPMED reference panel (version r2, N=194,512 haplotypes from N=97,256 individuals)^5^. Finally, all variants were submitted to the Michigan Imputation server (https://imputation.biodatacatalyst.nhlbi.nih.gov/). The server uses EAGLE (v2.4) to phase data and Minimac4 to perform genotype imputation to the reference panel. After quality control and genotype imputation of the genetic data, we kept only individuals of European ancestry (based on 1000Genomes clustering)^6^, and excluded individuals with a family relation (identity-by-descent ≥ 0.2)^7^, leaving 59 clinically diagnosed PSP-RS cases and 1513 controls for analysis.

### 3. Supplementary notes: Detecting potential clinical heterogeneity using genetic information

We leveraged the *MAPT* H1 haplotype, a well-known genetic risk factor for PSP^8^, to make inferences on misdiagnose rates in the Stage I clinical PSP dataset and its subgroups. We considered the confirmed pathological PSP cases as a reference of a pure PSP group, and the controls as a reference for a non-PSP population. Theoretically, for a mix of PSP and non-PSP samples, the expected value of the *MAPT* H1 allele frequency (AF) should range between its frequency in the pure/histopathological PSP (H1_Freq_=0.9199), and the non-PSP (H1_Freq_=0.7024) populations in our study, and should be proportional to the ratio of PSP/non-PSP samples in the mix. Thus, the *MAPT* H1 AF in the mixed sample would be equal to the *MAPT* H1 AF in the histopathological PSP sample minus the difference in AF between the histopathological PSP and control groups, (i.e. the total AF range between the pure PSP and non-PSP groups), multiplied by the proportion of misdiagnosed samples in the mix:

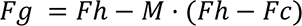

***Fg:*** *MAPT H1 AF in the mixed group; **Fh:** MAPT H1 AF in the histopathological PSP group (AF=0.9199); **M:** misdiagnose rate; **Fc:** MAPT H1 AF in the non PSP group (AF=0,7024); In the case of using logistic regression-derived effect sizes instead of frequencies, the log(OR) of these groups can be introduced instead of AF values*.

Rearranging the terms in this equation, we can estimate the percentage of misclassified samples in each group:

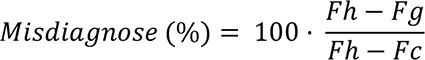

(*Fh* − *Fg*)*, difference in MAPT H1 AF between the histopathological PSP and mixed group*.

(*Fh* − *Fc*)*, difference in MAPT H1 AF between the histopathological PSP and the general population*.

The same logic can be applied using linear effect size estimates (log odds ratios) derived from logistic regressions instead of AFs. As population microstructure might affect the *MAPT* H1 allele frequency in the Iberian population^9^, we also calculated the misclassification rates using the log odds ratios obtained from fitting logistic regressions adjusted by the first four PCs in each subgroup. For better comparability between all groups, we included the controls from the discovery and validation setup (N=1,450) in all models. Additionally, because a proportion of the misclassified patients are expected to have Parkinson’s Disease (PD)^10^ and the MAPT H1 locus is known to also be a risk factor for PD, we used a OR=1.00 (neutral) and a OR=1.32 (Parkinson’s Disease MAPT H1)^11^ to calculate the effect size-based misdiagnosis estimations.

### 4. Supplementary notes: PRS calculation

We calculated the PRS of AD, PD, amyotrophic lateral sclerosis (ALS), PSP and all stroke (AS) using reported genome-wide significant variants^12–16^. We constructed the PRSs selecting only those variants with MAF>0.01 and imputation quality R2>0.3 in our dataset (Supplementary Tables 3-7). A few considerations were taken in the AD PRS calculation: a) No *APOE* variants were included, b) variants with MAF<0.01 were kept since these were previously validated in the original publication and they had very good imputation quality in our dataset (R2>0.9), c) because some controls of our study were included in the first stage of the AD GWAS, we used the independent effects reported exclusively in the second stage. To calculate the different PRSs, we designed a custom software (https://github.com/Pablo-GarGon/PRS_Generator) that added the dosages of the risk alleles multiplied by their reported effect sizes (betas). Finally, we also calculated the PRSs excluding the chromosome 17 to avoid pleiotropic effects caused by including variants linked to the *MAPT locus*.

### 5. Supplementary notes: A Decreased burden of known PSP genetic risk factors was detected in atypical PSP clinically diagnosed cases

Because, to our very best knowledge, this the first case-control PSP GWAS including clinical cases diagnosed using the recently proposed MDS criteria for probable PSP^17^, and our *MAPT* H1 association results suggested the existence of phenotypic heterogeneity in our clinical PSP cohort (Fig. 1), we decided to estimate genetically-based misclassification rates in the available Stage I clinical PSP subjects (Supplementary Table 1). We classified these samples into four subgroups based on two criteria: a) Samples with or without confirmed Richardson’s Syndrome and b) samples recruited by movement disorder units or memory clinics. The first classification allowed us to examine the specificity of the MDS criteria for assigning a probable PSP diagnosis to non-Richardson PSP (nR-PSP) patients, while the second one is useful to study potential ascertainment bias caused by the recruitment in different neurology set-ups (movement disorders units and memory clinics). As expected, the highest *MAPT* H1 frequency and effect size was found in the histopathological PSP group, and the misclassification estimates were considerably larger for nR-PSP than for PSP-RS samples (Fig. 1, Supplementary Table 1). Moreover, *MAPT* H1 frequencies were significantly reduced in the nR-PSP patients compared to histopathological PSP (χ²=6.29; DF=1; *p=*0.01), while no significant differences were observed for the PSP-RS patients in this comparison (χ²=0.16; DF=1; *p=*0.69), supporting the presence of misclassified patients in the nR-PSP group. Interestingly, PSP-RS samples had similar misclassification rates despite recruitment centre, while nR-PSP samples recruited in memory clinics displayed lower misdiagnose rates than those recruited by movement disorder units, although these differences were not significant (χ²=1.38; DF=1; *p=*0.24).

To further complement these results, we generated the AD, PD, ALS, PSP and AS PRSs using known disease susceptibility variants and compared the PRS values between our PSP subgroups and controls (Supplementary Fig. 1, Supplementary Table 10). The AD PRS was significantly increased in both nR-PSP subgroups: memory-clinic (*OR[95%CI]*=2.42[1.10–5.35]; *p=*0.03) and movement disorder unit-recruited (*OR[95%CI]*=2.64[1.13–6.22]; *p=*0.03). Meanwhile, the PD PRS was associated with disease status in both groups recruited by movement disorder units: PSP-RS (*OR[95%CI]*=1.90[1.22–2.96]; *p=*0.004) and nR-PSP (*OR[95%CI]*=1.72[1.00–2.93]; *p=*0.05). Finally, the PSP PRS was associated with disease status in all subgroups except movement disorder recruited nR-PSP (*OR[95%CI]*=1.20[0.96-1.52]; *p=*0.12), and no significant associations were found between disease status and the ALS and AS PRSs. Because *MAPT* is a pleiotropic risk factor for multiple neurodegenerative diseases, we repeated this analysis excluding chromosome 17. After removing variants in the *MAPT* locus, an increase could still be observed for the AD PRS in the nR-PSP subgroups and for the PD PRS in movement disorder unit-recruited subgroups, but the statistical significance was lost (Supplementary Fig. 1, Supplementary Table 7). Interestingly, the association with the PSP PRS was lost in both nR-PSP subgroups, and even showed a non-significant opposite effect direction in movement disorder recruited nR-PSP patients (*OR[95%CI]*=0.69[0.32-1.48]; *p=*0.34). Furthermore, none of the individual known PSP risk variants were associated with disease status in this subgroup either (Fig. 1). Our results indicate that the movement disorder recruited nR-PSP patients lacked an enrichment not only for *MAPT* H1 haplotypes but also for other known PSP risk factors. Subgroup-specific association of each individual risk variant constituting the different PRSs is available (Supplementary Tables 10-14).

Our data suggests two possible sources of contamination in our clinical dataset. On the one hand, nR-PSP patients who do not evolve to PSP-RS, the hallmark PSP phenotype, are hard to conclusively diagnose pre-mortem^18^. Thus, a weaker enrichment in known genetic PSP risk factors and, conversely, an enrichment in risk factors for other neurodegenerative diseases can be somewhat expected in the nR-PSP group. On the other hand, the observed enrichment in PD genetic risk variants in the context of movement disorder units suggests the presence of a fraction of misclassified parkinsonian PSP-like phenocopies in this clinical setting^19^. PSP-P patients comprise up to a third of the total number of PSP patients and are particularly challenging to accurately diagnose based solely on clinical symptoms^20^. As these patients are characterized by an initial phase with PD-compatible features, a higher chance of recruitment by movement disorder units rather than by memory clinics might explain our results^21^. However, a smaller risk effect of the *MAPT* H1 haplotype has also been described in PSP-P subjects compared to PSP-RS^22^, which may also explain the decrease in *MAPT* H1 frequency we observed in the nR-PSP groups. Overall, our findings suggest that there might be room for improvement of the specificity of the MDS criteria by increasing the diagnostic accuracy of atypical PSP phenotypes, specially PSP-P. Various biomarkers are under development to further help discriminate PSP from PSP-like patients: Real time quaking induced conversion (RT-QuIC) techniques to discard α-synuclein parkinsonian pathology, or for confirmation of 4R tauopathy are underway as CSF biomarkers, and MRI parameters like the magnetic resonance parkinsonism index are showing promising results in predicting PSP-RS conversion in early PSP-P subjects^21^.

### 6. Supplementary notes: Sensitivity analysis of PSP sentinels

Based on our previous results, we decided to run a sensitivity analysis for known PSP risk variants, based on GWS hits by Sanchez-Contreras *et al*^15^, stratifying our Stage I cases in three groups of decreasing PSP certainty: histopathological PSP, PSP-RS and nR-PSP. To improve comparability of the calculated estimates, we included the complete set of Stage I controls (N=1450) in all three strata. For all tested variants, we found consistent effect directions between the histopathological and PSP-RS groups (Supplementary Fig. 2). The effects in the nR-PSP group were smaller or even had opposite directions for the tested variants, except for rs242557, the variant tagging the *MAPT* H1c haplotype suggesting it may similarly impact genetic susceptibility to PSP-like syndromes. Finally, we decided to repeat the meta-analysis of the Stage I discovery and validation datasets, this time keeping only the PSP-RS samples in the validation set. After removing nR-PSP samples, the effect sizes were stronger in all PSP susceptibility variants (Supplementary Table 9), showcasing the benefits of removing these samples with lower diagnostic accuracy from our GWAS analysis.

### 7. Supplementary notes: Enrichment analysis

Enrichment analysis was performed using WebGestalt (https://www.webgestalt.org/) over-representation analysis^23^. Briefly, this method evaluates the fraction of genes which constitute the different gene sets that are present in the list of genes introduced, testing the significance using a 2x2 contingency table^24^. We introduced the list of replicated genes (*MAPT, MOBP, EIF2AK3, STX6, DUSP10, SLCO1A2*) and our novel *NFASC* gene and ran the over representation analysis algorithm, enriching specifically for non-redundant gene ontology terms and using the “Affy Axiom Biobank1” option as the reference gene set. Although none of the top enriched terms were significant after multiple test correction (Extended Data Table 6), the term GO:0097386, representing glial cell projection was borderline significant (FDR adjusted p-value = 0.06) and several of the top ranked terms were related to axon ensheathment an d myelination, pinpointing these processes as those more represented by our list of genes.

### 8. Supplementary notes: Gene co-expression network analysis

Gene co-expression networks were generated using GeneFriends software (https://www.genefriends.org/)25, including all interesting confirmed loci and candidates (*MAPT, MOBP, EIF2AK3, STX6, DUSP10, SLCO1A2, NFASC* and *CNTN2)* using the Sequence Read Archive (SRA) human tissue co-expression map as a reference, which comprises 20 tissues from 46080 RNA-seq samples. Briefly, this method first estimates correlations between each pair of genes, then uses co-expression associations to construct the co-expression network, and finally modules, i.e. groups of co-expressed genes are identified using clustering techniques, allowing to group those genes with similar expression patterns across the different samples. The Pearson correlation threshold was adjusted to 0.6 to allow us to identify the top genes co-expressed with our main cluster, consisting of *MOBP*, *SLCO1A2* and *NFASC*. We found that *MARK1*, as well as three additional lncRNAs and one pseudogene, were strongly co-expressed (R>0.6) with this gene cluster (Extended Data Fig. 5).

### Supplementary Table Legends

**Supplementary Table 1.** QC summary of contributed samples by collaborating center and stage.

**Supplementary Table 2.** Age linear regressions results for the complete set of controls used in PSP/DEGESCO Stage I (N=1450). Genomic coordinates correspond to the GRCh38 assembly.

**Supplementary Table 3.** Reported summary statistics of the variants used to construct the Alzheimer’s Disease (AD) PRS. We used exclusively effects from stage 2 of the Bellenguez *et al.* study^12^. Genomic coordinates correspond to the GRCh38 assembly.

**Supplementary Table 4.** List of the variants used to construct the amyotrophic lateral sclerosis (ALS) PRS. These summary statistics correspond to the European ancestry GWAS from the original study. Genomic coordinates correspond to the GRCh38 assembly.

**Supplementary Table 5.** List of the variants used to construct the all stroke (AS) PRS. Genomic coordinates correspond to the GRCh38 assembly.

**Supplementary Table 6.** List of the variants used to construct the Parkinson’s Disease (PD) PRS. Genomic coordinates correspond to the GRCh38 assembly.

**Supplementary Table 7.** List of the variants used to construct the progressive supranuclear palsy (PSP) PRS. Genomic coordinates correspond to the GRCh38 assembly.

**Supplementary Table 8.** Estimation of the percentage of misdiagnosed clinical PSP samples classified by those with/without Richardson’s Syndrome and by recruitment centre (Movement Disorder Unit/Memory Clinic) in PSP/DEGESCO Stage I. ORs were calculated fitting logistic regressions adjusted by the first four PCs and using the same set of controls in all groups (N=1450). Percentage of misdiagnosed samples was calculated based on MAPT H1 haplotype frequencies and log odds ratios. Age data represents age at death, age at onset of motor symptoms and age at last visit for histopathological PSP, clinical PSP and controls, respectively.

**Supplementary Table 9.** Sensitivity analysis results for the replicated variants. We showcase the results obtained in the previous study and the PSP/DEGESCO meta-analysis results including and excluding nR-PSP samples. Models were adjusted by the first four PCs. EA, effect allele; OA, other allele; OR, odds ratio; P, p-value.

**Supplementary Table 10.** Association in case control logistic regression of neurodegenerative disease PRSs with PSP in the different clinical subgroups. PRSs were calculated using all reported variants, and removing variants in chr17 to account for MAPT pleiotropy. Associations are reported in the following format: OR [95%CI]; P. OR, odds ratio; PRS, polygenic risk score; AD, Alzheimer’s Disease; PD, Parkinson’s Disease; ALS, amyotrofic lateral sclerosis; PSP, progressive supranuclear palsy; AS, all stroke.

**Supplementary Table 11.** Association in case control logistic regression of the individual variants used to construct the Alzheimer’s Disease (AD) PRS with the diferent PSP subgroups in PSP/DEGESCO Stage I.

**Supplementary Table 12.** Association in case control logistic regression of the individual variants used to construct the amyotrophic lateral sclerosis (ALS) PRS with the diferent PSP subgroups in PSP/DEGESCO Stage I.

**Supplementary Table 13.** Association in case control logistic regression of the individual variants used to construct the all stroke (AS) PRS with the diferent PSP subgroups in PSP/DEGESCO Stage I.

**A Supplementary Table 14.** Association in case control logistic regression of the individual variants used to construct the Parkinson’s disease (PD) PRS with the diferent PSP subgroups in PSP/DEGESCO Stage I.

**Supplementary Table 15.** Association in case control logistic regression of the individual variants used to construct the progressive supranuclear palsy (PSP) PRS with the diferent PSP subgroups in PSP/DEGESCO Stage I.

### Supplementary Figures

**Supplementary Fig. 1.**
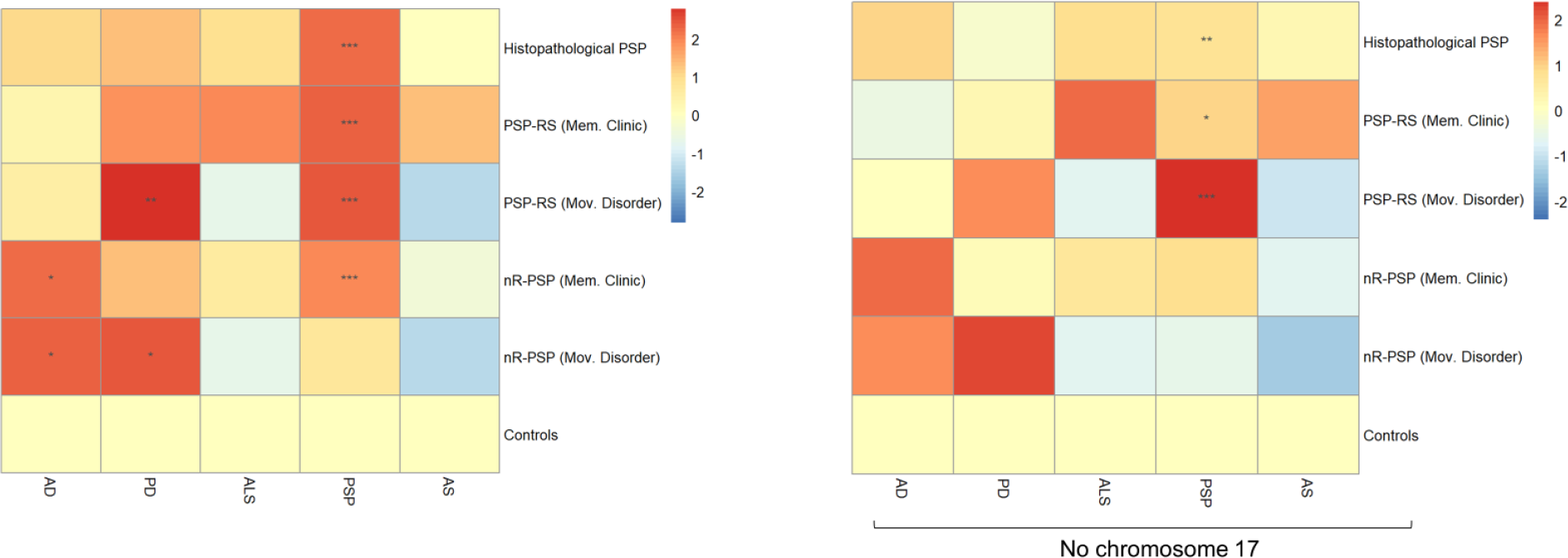
Mean AD, PD, ALS, PSP and AS PRS in each PSP subgroup including and excluding variants in chr17. Values were zero-centred with respect to the control group to better reflect the increase/decrease in mean PRS in the PSP groups with respect to the controls. OR, odds ratio; PRS, polygenic risk score; AD, Alzheimer’s Disease; PD, Parkinson’s Disease; ALS, amyotrophic lateral sclerosis; PSP, progressive supranuclear palsy; AS, all stroke; *** p<0.001; ** p<0.01; * p<0.05.

**Supplementary Fig. 2.**
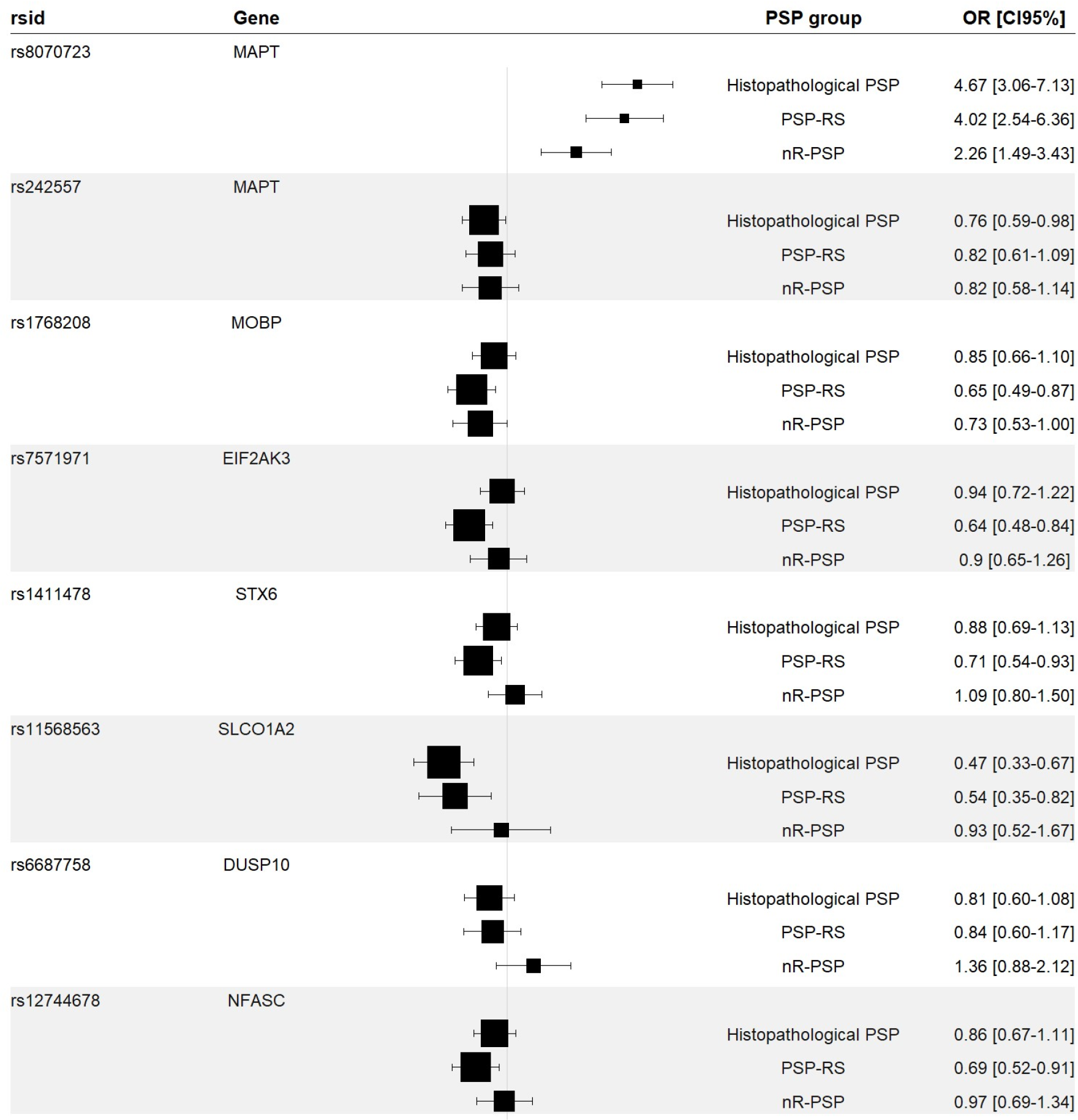
Forest plot displaying effect sizes of SNPs in the histopathological and clinical groups (PSP-RS or nR-PSP) of PSP/DEGESCO Stage I. Models were adjusted by PCs1-4 and included the same set of controls (N=1450) to improve comparability. OR, odds ratio; CI95%, 95% confidence interval.

## References

1. Boxer, A. L. et al. Advances in progressive supranuclear palsy: new diagnostic criteria, biomarkers, and therapeutic approaches. The Lancet Neurology 16, 552–563 (2017).

2. Baker, M. et al. Association of an Extended Haplotype in the Tau Gene with Progressive Supranuclear Palsy. Human Molecular Genetics 8, 711–715 (1999).

3. PSP Genetics Study Group et al. Identification of common variants influencing risk of the tauopathy progressive supranuclear palsy. Nat Genet 43, 699–705 (2011).

4. Sanchez-Contreras, M. Y. et al. Replication of progressive supranuclear palsy genome-wide association study identifies SLCO1A2 and DUSP10 as new susceptibility loci. Mol Neurodegeneration 13, 37 (2018).

5. Chen, J. A. et al. Joint genome-wide association study of progressive supranuclear palsy identifies novel susceptibility loci and genetic correlation to neurodegenerative diseases. Mol Neurodegeneration 13, 41 (2018).

6. Rojas, I. de et al. Common variants in Alzheimer’s disease and risk stratification by polygenic risk scores. Nature Communications 12, 3417 (2021).

7. Taliun, D. et al. Sequencing of 53,831 diverse genomes from the NHLBI TOPMed Program. Nature 590, 290–299 (2021).

8. Höglinger, G. U. et al. Clinical diagnosis of progressive supranuclear palsy: The movement disorder society criteria: MDS Clinical Diagnostic Criteria for PSP. Mov Disord. 32, 853– 864 (2017).

9. Escott-Price, V. & Hardy, J. Genome-wide association studies for Alzheimer’s disease: bigger is not always better. Brain Communications 4, fcac125 (2022).

10. Wang, H. et al. Whole-Genome Sequencing Analysis Reveals New Susceptibility Loci and Structural Variants Associated with Progressive Supranuclear Palsy. Preprint at http://medrxiv.org/lookup/doi/10.1101/2023.12.28.23300612 (2023).

11. Farrell, K. et al. Genetic, Transcriptomic, Histological, and Biochemical Analysis of Progressive Supranuclear Palsy Implicates Glial Activation and Novel Risk Genes. Preprint at http://biorxiv.org/lookup/doi/10.1101/2023.11.09.565552 (2023).

12. Sherman, D. L. et al. Neurofascins Are Required to Establish Axonal Domains for Saltatory Conduction. Neuron 48, 737–742 (2005).

13. Zhang, A. et al. Neurofascin 140 Is an Embryonic Neuronal Neurofascin Isoform That Promotes the Assembly of the Node of Ranvier. J. Neurosci. 35, 2246–2254 (2015).

14. Kira, J., Yamasaki, R. & Ogata, H. Anti-neurofascin autoantibody and demyelination. Neurochemistry International 130, 104360 (2019).

15. Yang, C. et al. Genomic atlas of the proteome from brain, CSF and plasma prioritizes proteins implicated in neurological disorders. Nat Neurosci 24, 1302–1312 (2021).

16. Zoupi, L. et al. The function of contactin-2/TAG-1 in oligodendrocytes in health and demyelinating pathology. Glia 66, 576–591 (2018).

17. Sjöstedt, E. et al. An atlas of the protein-coding genes in the human, pig, and mouse brain. Science 367, eaay5947 (2020).

18. Gobert, R. P. et al. Convergent functional genomics of oligodendrocyte differentiation identifies multiple autoinhibitory signaling circuits. Mol Cell Biol 29, 1538–1553 (2009).

19. Gu, G. J. et al. Role of Individual MARK Isoforms in Phosphorylation of Tau at Ser262 in Alzheimer’s Disease. Neuromol Med 15, 458–469 (2013).

20. Kahlson, M. A. & Colodner, K. J. Glial Tau Pathology in Tauopathies: Functional Consequences. J Exp Neurosci 9s2, JEN.S25515 (2015).

21. Allen, M. et al. Conserved brain myelination networks are altered in Alzheimer’s and other neurodegenerative diseases. Alzheimer’s & Dementia 14, 352–366 (2018).

22. Briel, N., Pratsch, K., Roeber, S., Arzberger, T. & Herms, J. Contribution of the astrocytic tau pathology to synapse loss in progressive supranuclear palsy and corticobasal degeneration. Brain Pathology 31, e12914 (2021).

23. Ling, H. et al. Astrogliopathy predominates the earliest stage of corticobasal degeneration pathology. Brain 139, 3237–3252 (2016).

24. Graham, S. E. et al. The power of genetic diversity in genome-wide association studies of lipids. Nature 600, 675–679 (2021).

## References

25. Hauw, J.-J. et al. Preliminary NINDS neuropathologic criteria for Steele-Richardson-Olszewski syndrome (progressive supranuclear palsy). Neurology 44, 2015–2015 (1994).

26. Van Der Flier, W. M. & Scheltens, P. Amsterdam Dementia Cohort: Performing Research to Optimize Care. JAD 62, 1091–1111 (2018).

27. Konijnenberg, E. et al. The EMIF-AD PreclinAD study: study design and baseline cohort overview. Alz Res Therapy 10, 75 (2018).

28. Holstege, H. et al. The 100-plus Study of cognitively healthy centenarians: rationale, design and cohort description. Eur J Epidemiol 33, 1229–1249 (2018).

29. Jansen, I. E. et al. Genome-wide meta-analysis for Alzheimer’s disease cerebrospinal fluid biomarkers. Acta Neuropathol 144, 821–842 (2022).

30. Moreno-Grau, S. et al. Genome-wide association analysis of dementia and its clinical endophenotypes reveal novel loci associated with Alzheimer’s disease and three causality networks: The GR@ACE project. Alzheimer’s & Dementia 15, 1333–1347 (2019).

31. Gogarten, S. M. et al. Genetic association testing using the GENESIS R/Bioconductor package. Bioinformatics 35, 5346–5348 (2019).

32. Das, S. et al. Next-generation genotype imputation service and methods. Nat Genet 48, 1284–1287 (2016).

33. The 1000 Genomes Project Consortium et al. A global reference for human genetic variation. Nature 526, 68–74 (2015).

34. Anderson, C. A. et al. Data quality control in genetic case-control association studies. Nature Protocols 5, 1564–1573 (2011).

35. Taylor, J. M. Choosing the number of controls in a matched case-control study, some sample size, power and efficiency considerations. Stat Med 5, 29–36 (1986).

36. Baker, E. et al. What does heritability of Alzheimer’s disease represent? PLoS ONE 18, e0281440 (2023).

37. Sánchez-Juan, P. et al. The MAPT H1 Haplotype Is a Risk Factor for Alzheimer’s Disease in APOE ε4 Non-carriers. Front. Aging Neurosci. 11, 327 (2019).

38. Willer, C. J., Li, Y. & Abecasis, G. R. METAL: fast and efficient meta-analysis of genomewide association scans. Bioinformatics 26, 2190–2191 (2010).

39. Balduzzi, S., Rücker, G. & Schwarzer, G. How to perform a meta-analysis with R: a practical tutorial. Evid Based Mental Health 22, 153–160 (2019).

40. Wang, J., Vasaikar, S., Shi, Z., Greer, M. & Zhang, B. WebGestalt 2017: a more comprehensive, powerful, flexible and interactive gene set enrichment analysis toolkit. Nucleic Acids Research 45, W130–W137 (2017).

41. Raina, P. et al. GeneFriends: gene co-expression databases and tools for humans and model organisms. Nucleic Acids Research 51, D145–D158 (2023).

## References

1. Sanchez-Contreras, M. Y. et al. Replication of progressive supranuclear palsy genome-wide association study identifies SLCO1A2 and DUSP10 as new susceptibility loci. Mol Neurodegeneration 13, 37 (2018).

2. Chen, J. A. et al. Joint genome-wide association study of progressive supranuclear palsy identifies novel susceptibility loci and genetic correlation to neurodegenerative diseases. Mol Neurodegeneration 13, 41 (2018).

3. Wang, H. et al. Whole-Genome Sequencing Analysis Reveals New Susceptibility Loci and Structural Variants Associated with Progressive Supranuclear Palsy. Preprint at http://medrxiv.org/lookup/doi/10.1101/2023.12.28.23300612 (2023).

4. Farrell, K. et al. Genetic, Transcriptomic, Histological, and Biochemical Analysis of Progressive Supranuclear Palsy Implicates Glial Activation and Novel Risk Genes. Preprint at http://biorxiv.org/lookup/doi/10.1101/2023.11.09.565552 (2023).

5. PSP Genetics Study Group et al. Identification of common variants influencing risk of the tauopathy progressive supranuclear palsy. Nat Genet 43, 699–705 (2011).

6. The 1000 Genomes Project Consortium et al. A global reference for human genetic variation. Nature 526, 68–74 (2015).

7. Yang, C. et al. Genomic atlas of the proteome from brain, CSF and plasma prioritizes proteins implicated in neurological disorders. Nat Neurosci 24, 1302–1312 (2021).

8. Karlsson, M. et al. A single–cell type transcriptomics map of human tissues. Sci. Adv. 7, eabh2169 (2021).

9. Wang, J., Vasaikar, S., Shi, Z., Greer, M. & Zhang, B. WebGestalt 2017: a more comprehensive, powerful, flexible and interactive gene set enrichment analysis toolkit. Nucleic Acids Research 45, W130–W137 (2017).

10. Raina, P. et al. GeneFriends: gene co-expression databases and tools for humans and model organisms. Nucleic Acids Research 51, D145–D158 (2023).

## Supplementary References

1. de Rojas, I. et al. Common variants in Alzheimer’s disease and risk stratification by polygenic risk scores. Nature Communications (2021) doi:10.1038/s41467-021-22491-8.

2. Moreno-Grau, S. et al. Genome-wide association analysis of dementia and its clinical endophenotypes reveal novel loci associated with Alzheimer’s disease and three causality networks: The GR@ACE project. Alzheimer’s and Dementia (2019) doi:10.1016/j.jalz.2019.06.4950.

3. Gogarten, S. M. et al. Genetic association testing using the GENESIS R/Bioconductor package. Bioinformatics (2019) doi:10.1093/bioinformatics/btz567.

4. Das, S. et al. Next-generation genotype imputation service and methods. Nat Genet 48, 1284–1287 (2016).

5. Taliun, D. et al. Sequencing of 53,831 diverse genomes from the NHLBI TOPMed Program. Nature 590, 290–299 (2021).

7. Anderson, C. A. et al. Europe PMC Funders Group Data quality control in genetic case-control association studies. Nature Protocols (2011) doi:10.1038/nprot.2010.116.Data.

8. Baker, M. et al. Association of an Extended Haplotype in the Tau Gene with Progressive Supranuclear Palsy. Human Molecular Genetics 8, 711–715 (1999).

9. Gayán, J. et al. Genetic Structure of the Spanish Population. BMC Genomics 11, 326 (2010).

10. Höglinger, G. U. et al. Identification of common variants influencing risk of the tauopathy progressive supranuclear palsy. in Nature Genetics (2011). doi:10.1038/ng.859.

11. Simón-Sánchez, J. et al. Genome-wide association study reveals genetic risk underlying Parkinson’s disease. Nat Genet 41, 1308–1312 (2009).

12. Bellenguez, C. et al. New insights into the genetic etiology of Alzheimer’s disease and related dementias. Nat Genet 54, 412–436 (2022).

13. Nalls, M. A. et al. Identification of novel risk loci, causal insights, and heritable risk for Parkinson’s disease: a meta-analysis of genome-wide association studies. Lancet Neurol 18, 1091–1102 (2019).

14. van Rheenen, W. et al. Common and rare variant association analyses in amyotrophic lateral sclerosis identify 15 risk loci with distinct genetic architectures and neuron-specific biology. Nat Genet 53, 1636–1648 (2021).

15. Sanchez-Contreras, M. Y. et al. Replication of progressive supranuclear palsy genome-wide association study identifies SLCO1A2 and DUSP10 as new susceptibility loci. Mol Neurodegeneration 13, 37 (2018).

16. Mishra, A. et al. Stroke genetics informs drug discovery and risk prediction across ancestries. Nature 611, 115–123 (2022).

17. Höglinger, G. U. et al. Clinical diagnosis of progressive supranuclear palsy: The movement disorder society criteria: MDS Clinical Diagnostic Criteria for PSP. Mov Disord. 32, 853–864 (2017).

18. Respondek, G. et al. Accuracy of the national institute for neurological disorders and stroke/society for progressive supranuclear palsy and neuroprotection and natural history in Parkinson plus syndromes criteria for the diagnosis of progressive supranuclear palsy. Movement Disorders 28, 504–509 (2013).

19. Boxer, A. L. et al. Advances in progressive supranuclear palsy: new diagnostic criteria, biomarkers, and therapeutic approaches. The Lancet Neurology 16, 552–563 (2017).

20. Alster, P., Madetko, N., Koziorowski, D. & Friedman, A. Progressive Supranuclear Palsy—Parkinsonism Predominant (PSP-P)—A Clinical Challenge at the Boundaries of PSP and Parkinson’s Disease (PD). Front. Neurol. 11, 180 (2020).

21. Coughlin, D. G. & Litvan, I. Progressive supranuclear palsy: Advances in diagnosis and management. Parkinsonism Relat Disord 73, 105–116 (2020).

22. Williams, D. R. et al. Characteristics of two distinct clinical phenotypes in pathologically proven progressive supranuclear palsy: Richardson’s syndrome and PSP-parkinsonism. Brain 128, 1247–1258 (2005).

23. Wang, J., Vasaikar, S., Shi, Z., Greer, M. & Zhang, B. WebGestalt 2017: a more comprehensive, powerful, flexible and interactive gene set enrichment analysis toolkit. Nucleic Acids Research 45, W130–W137 (2017).

24. Khatri, P., Sirota, M. & Butte, A. J. Ten Years of Pathway Analysis: Current Approaches and Outstanding Challenges. PLoS Comput Biol 8, e1002375 (2012).

25. Raina, P. et al. GeneFriends: gene co-expression databases and tools for humans and model organisms. Nucleic Acids Research 51, D145–D158 (2023).

